# Improving patient flow through hospitals with machine learning based discharge prediction

**DOI:** 10.1101/2023.05.02.23289403

**Authors:** Jia Wei, Jiandong Zhou, Zizheng Zhang, Kevin Yuan, Qingze Gu, Augustine Luk, Andrew J Brent, David A. Clifton, A. Sarah Walker, David W. Eyre

## Abstract

Accurate predictions of hospital discharge events could help improve patient flow through hospitals and the efficiency of care delivery. However, the potential of integrating machine learning with diverse electronic health records (EHR) data for this task has not been fully explored. We used EHR data from 01 February 2017 to 31 January 2020 in Oxfordshire, UK to predict hospital discharges in the next 24 hours. We fitted separate extreme gradient boosting models for elective and emergency admissions, trained using the first two years of data and tested using the final year of data. We examined individual-level and hospital-level model performance and evaluated the impact of training data size and recency, prediction time of day, and performance in different subgroups. Our individual patient level models for elective and emergency admissions achieved AUCs of 0.87 and 0.86, AUPRCs of 0.66 and 0.64 and F1 scores of 0.61 and 0.59, respectively, substantially better than a baseline logistic regression model. Aggregating individual probabilities, the total daily number of hospital discharges could also be accurately estimated, with mean absolute errors of 8.9% (elective admissions) and 4.9% (emergency admissions). The most informative predictors included antibiotic prescriptions, other medications, and hospital capacity factors. Performance was generally robust across patient subgroups and different training strategies, but lower in patients with longer lengths of stay and those who eventually died in hospital. Our findings highlight the potential of machine learning in optimising hospital patient flow and facilitating patient care and recovery.

## Introduction

Increasing demand for healthcare, driven by changing population demographics, a rise in the prevalence of chronic diseases, societal changes, and technological advances, places significant strain on hospital resources^1^. In the United Kingdom (UK), the National Health Service (NHS) has faced escalating demand pressures in recent years, with an increasing number of admissions, prolonged waiting times in Emergency Departments, and financial challenges^2,3^. This has been exacerbated by the COVID-19 pandemic, resulting in substantial backlogs in both urgent and routine care^4^. With healthcare resources being inherently limited, there is a pressing need to enhance the efficiency of healthcare services and improve hospital capacity management. A critical component is patient flow within hospitals, referring to the movement of patients from admission to discharge while ensuring they receive appropriate care and resources^5^. Optimising this could improve patient experiences, avoid delays in treatment, improve health outcomes, and reduce costs^6^.

Accurately predicting when patients will be discharged from the hospital could improve patient flow, e.g. prompting clinicians when patients are approaching readiness for discharge, facilitating booking transport home, enabling timely preparation of discharge medication and documentation, and pre-emptively arranging room cleaning. Currently, discharge predictions are made in most hospitals at the individual patient level by clinical teams based on the patient’s diagnosis and status and are updated throughout their hospital stay. However, these assessments can be subjective and variable and may not always be captured in electronic healthcare record (EHR) systems, posing challenges to efficient operational management. Therefore, there is growing interest in leveraging automated prediction models to forecast the length of stay (LOS) and discharge timing, both individually and hospital-wide.

Discharge prediction has therefore become a key target for clinical machine learning researchers. Several previous studies have attempted to predict discharge within a fixed time window (**Table 1**), with studies typically predicting discharge within the next 24, 48, or 72 hours. Some of these studies have focused on specific patient populations, e.g. surgical patients^7,8^ or those with cardiovascular disease^9^, while others predict discharge for whole hospitals^10–14^. A range of different classical machine learning approaches have been evaluated, including random forests, gradient boosted trees, and multilayer perceptron neural networks. Input features being considered typically include details relating to the index date the prediction is being made on, past medical history, prior length of stay, demographics, current vital signs and laboratory markers, diagnoses, procedures, and medications. Performance is typically modest in most models, although some perform better, with area under the receiver operating curve (AUC) values ranging from 0.70-0.86 in whole hospital models. One notable exception is a model that also included data on EHR-user interactions^10^, such as the frequency of clinical notes being updated, viewed, or printed, which achieved an AUC of 0.92 for predicting discharge within 24 hours. However, this study only included the first admission for each patient, potentially under-representing complex patients who may be frequently re-admitted. It also excluded patients who died during hospitalisation. Additionally, in most previous studies several key areas relevant to implementation were not fully explored, including the impact of training data size and recency, the time of day or day during the week a prediction is made, and performance relative to length of stay and specific patient subgroups, including those affected by health inequalities. Most studies either evaluated individual-level discharge prediction performance or hospital-wide predictions, but did not combine the two in a single approach.

**Table 1.** Previous studies predicting discharge within a fixed time window. We searched Google Scholar and PubMed for studies up to 30 April 2024, using the search terms ‘machine learning’ AND (‘hospital discharge prediction’, OR ‘patient flow’).

| Publication/Year | Population | Outcome | Model | Features | Performance, AUC where available |
| --- | --- | --- | --- | --- | --- |
| Ahn et al. 2021 | Inpatients with cardiovascular diseases admitted to Asan Medical Centre in Korea between 2000 to 2016; 669,667 records | Discharge within the next 72 hours (predictions not made on the day of discharge) | Extreme gradient boosting (XGB) | Index date-related features, diagnosis, operations, medications, procedures, laboratory tests, past medical history (last 3 years) | AUC 0.87 |
| Zhang et al. 2021 | Adult patients admitted to Vanderbilt University Medical Centre in 2019; 26,283 patients | Discharge within 24 hours | Light gradient boosting machine (LGBM) | Age, race, gender, insurance, user-EHR interactions (e.g. view/modify/export EHR entries), past medical history (Phecodes), discharge units, length of stay, discharge time, discharge day of week | AUC 0.92 with user-EHR interactions<br>AUC 0.86 without user-EHR interactions. |
| Barnes et al. 2016 | Patients admitted to a mid-Atlantic academic medical centre from 2011-2013; 8,852 patient visits and 20,243 individual patient days | Discharge within 7 and 17 hours (from 7 am). | Logistic regression (LR); Random Forest (RF) | Gender, ethnicity, age, insurance, reason for visit, observation status, discharge location, patient census, day of week, elapsed length of stay | Sensitivity: LR: 65.9; RF: 60.0<br>Specificity: LR: 52.8; RF: 66.0 |
| Levin et al. 2021 | Adult patients admitted to a community hospital in Maryland, USA between April 2016 and August 2019; 120,780 | Discharge on the same day, by the next day, within the next 2 days | RF | Demographic, administrative, temporal, medication, other interventions, diagnostics, monitoring, rehabilitation, | AUC 0.80 (same day)<br>AUC 0.70 (next day) |
|  | discharge for 12,470 patients |  |  | consults, diet and more complex clinical markers (pain management, substance abuse, sepsis, cardiac arrest, acute kidney injury) |  |
| Bertsimas et al. 2021 | Inpatients admitted at Beth Israel Deaconess Medical Centre between January 2017 and August 2018; 63,432 unique admissions (41,726 unique patients) | Discharge within 1 day, discharge within 2 days | LR, CART decision trees, Optimal trees, RF, Gradient boosted trees | Diagnosis, medications, laboratory results, body mass index, type of diet, level of activity and autonomy, socioeconomic factors, operations, laboratory results, and vitals | Discharge within 1 day, AUC 0.84<br>Discharge within 2 days, AUC 0.82 |
| Safavi et al. 2019 | Adult patients discharged from inpatient surgical care in the US from May 1, 2016, to August 31, 2017; 15,201 hospital discharges | Discharge within 24 hours | Multilayer perceptron neural network | Demographics, surgery information, clinician orders, clinical test results, bedside assessments, clinical recommendations, medication administration, catheter information, care team notes | AUC 0.84 |
| Lazar et al. 2020 | Adult surgical patients discharged from inpatient care between July 2018 and February 2020; 10,904 patients during 12,493 inpatient visits | Discharge within 48 hours | RF | Age, sex, admission source, laboratory measurements, and vitals | AUC 0.81 |
| Ward et al. 2021 | Patient encounters from 14 different Kaiser Permanente facilities in northern California from November 1, 2015 to | Discharge within 1 day | LR, Lasso, RF, GBM | Age, gender, admit type, admission and hourly LAPS2 and COPS2 scores, diagnoses, hourly number of orders, medications, time | GBM, AUC 0.73 |
|  | December 31, 2017;<br>910,366 patient-days<br>across 243,696 patients<br>hospitalisations |  |  | since admission, do-not-<br>resuscitate or comfort care |  |

In this study, we used diverse EHR-derived features and data from a large UK teaching hospital group to develop machine learning models to predict individual patient-level hospital discharge within the next 24 hours. By aggregating individual predictions, our models were also successful in predicting the total hospital-wide number of discharges expected.

## Methods

### Data and setting

We used data from the Infections in Oxfordshire Research Database (IORD) which contains deidentified electronic healthcare records from Oxford University Hospitals (OUH) NHS Foundation Trust. OUH consists of four teaching hospitals in Oxfordshire, United Kingdom, with a total of ∼1100 beds, serving a population of ∼755,000 people and providing specialist services to the surrounding region.

We extracted data for all adult inpatients (≥16 years) from 01 February 2017 to 31 January 2020 who had an ordinary admission (i.e., excluding day case, regular day admission, and regular night admissions). We excluded patients whose admission specialty was obstetrics or paediatrics, as these specialties used a different EHR system and/or discharge pathway. We grouped admissions into elective admissions (those scheduled in advance) and emergency admissions (those who entered hospital through the Emergency Department or other emergency admissions units) based on admission method codes (**Figure S1**).

### Model features

Domain knowledge and prior literature were used to determine which features within the dataset were potentially informative for predicting patient discharge. Input features included index-date related features, patient demographics, comorbidities, current admission features, previous length of stay for patients on the same ward, current diagnosis, recent procedures, antibiotic prescriptions, other medications, microbiology results, radiology investigations, prior hospital stays and readmissions in the last year, hospital factors, vital signs measurements, and laboratory test results (**Table S1**, **Supplementary methods**).

### Prediction task

We predicted hospital discharge events within 24 hours of an index date and time for all patients currently in the hospital (individual-level predictions). Model prediction probabilities were also aggregated to predict the total number of patients currently in hospital who would be discharged within the next 24 hours following the index date time (hospital-level predictions). We used a pragmatic and operationally relevant endpoint, predicting discharges from hospital with any outcome (discharged alive, transferred to another hospital, or died).

Each patient contributed once to the dataset per day during their hospital admission. Separate models were built for emergency and elective admissions as predictive features may be different depending on the reason for admission. For the main analyses, predictions were made at 12 pm for both the training and test data, and the probability of a patient being discharged by 12 pm on the following day was obtained. However, in real-world use model predictions are likely to be applied throughout the day as new patients are admitted and others discharged. We therefore performed three sets of sensitivity analyses: 1) train and predict discharge at other times of day (midnight, 6 am, 6 pm); 2) train the model and predict discharge using data drawn randomly throughout the day, using data available at 2 hourly intervals, i.e., midnight, 2 am, 4 am, …, 10 pm; 3) train and predict discharge at different times of day (e.g. train at 12 pm, test at 6 am).

We used extreme gradient boosting (XGB) models to predict discharge within the next 24 hours from an index date. Models were trained using data from the first two years of the study (01 February 2017 to 31 January 2019) and evaluated using data from the final year of the study (01 February 2019 to 31 January 2020). To examine whether the trained model could be used to predict hospital discharge following the COVID-19 pandemic, we also used data from 01 February 2021 to 31 January 2022 as an additional held-out test dataset.

We randomly split the training data, using 80% of the data for the main model training. Within this, a Bayesian optimisation for hyperparameters was performed by employing Tree-based Parzen estimators (TPE) to search through a wide potential hyperparameter space, maximising the AUC during 5-fold cross-validation. Details of model tuning are provided in the **Supplementary methods**. We used the built-in scale_pos_weight in XGB classifier to account for class imbalance, i.e. the fact there are more non-discharge events than discharges. No imputation of missing data was performed, as XGB can handle missing data by design. We used the remaining 20% of the training data as validation data for feature selection, calibrating the predicted probability from XGB models using isotonic regression^15,16^ and determining the best threshold for predicting a discharge event by optimising the F1 score (jointly maximising precision and recall). Initially, all 1,152 features were used to fit each model. Models were then re-fitted with progressively fewer features, retaining the top-ranked features from each full model. Performance in the validation data and training time was used to select the optimal number and list of input features for the main analyses. The model pipeline is shown in **Figure 1**. Model performance was compared to a baseline logistic regression with fewer selected features (age, sex, day of the week, hours since admission), for benchmarking against a relatively simple model.

**Figure 1.**
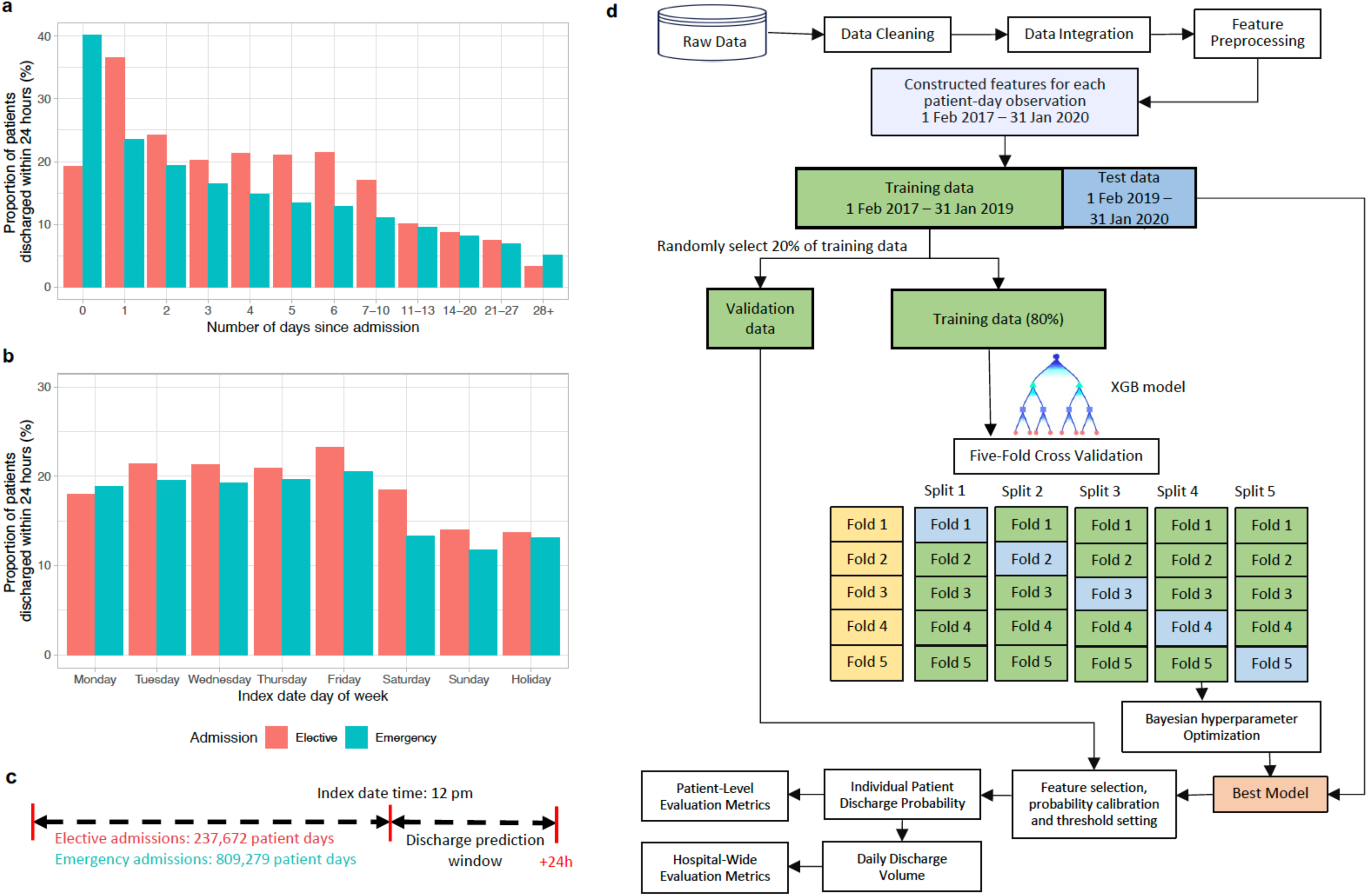
Overview of model development. **a)** The proportion of patients discharged from hospital within the next 24 hours in elective and emergency admissions by number of days since admission. **b)** The proportion of patients discharged from hospital within the next 24 hours in elective and emergency admissions by day of week of the index date. **c)** Diagram of the prediction problem. The binary prediction problem was defined by classifying the outcome as ‘positive’ (discharge occurred within the next 24 h) or ‘negative’ (discharge did not occur within the next 24h) separately for elective and emergency admissions. Predictions were made at 12 pm. **d)** Analysis pipeline for the prediction of hospital discharge within the next 24 hours. Extreme gradient boosting (XGB) models were trained on the extracted labels and features from admissions between 01 February 2017 to 31 January 2019, and was tested on admissions between 01 February 2019 and 31 January 2020. Five-fold cross validation was used for hyperparameter tuning, and 20% randomly selected validation data was used for feature selection, probability calibration, and threshold setting. The best model was used to predict hospital discharges in the test data, and model performance was examined.

### Performance assessment

Individual-level model performance was evaluated using sensitivity (recall), specificity, balanced accuracy (arithmetic mean of sensitivity and specificity), positive predictive value (PPV, precision), negative predictive value (NPV), F1 score (harmonic mean of precision and recall), AUC, and area under the precision-recall curve (AUPRC). PPV and NPV provide interpretable real-world metrics relating to actual discharge decision making and prediction performance. Although they depend on the prevalence of discharge events, our estimates are likely to apply to hospitals with similar daily discharge rates. F1 scores and AUPRC are also impacted by prevalence, and for a given sensitivity and specificity will be lower as prevalence falls, which should be considered when comparing subgroups with different discharge prevalence.

For hospital-level prediction, we summarised the accuracy of predictions of the total number of patients discharged using normalised mean absolute error (MAE, %), i.e. the mean of the differences in predicted and actual discharges each day (over the 365 predictions in the test dataset) divided by the mean number of discharges per day.

We calculated the SHapley Additive exPlanations (SHAP) values^17^ for each feature in the training dataset to determine feature importance.

We examined the model performance in different subgroups by age, sex, ethnicity, index of multiple deprivation score, weekday of the index date, admission specialty, comorbidity score, source of admission, days since admission, and discharge outcome (alive or died). We also used equalised odds differences to compare model fairness, by checking if either the per subgroup true positive rate or true negative rate differed from the overall rate by greater than an illustrative threshold of 0.1^18^.

We also evaluated model performance using the same test data, but with different lengths of training data from 1 to 24 months, and the impact of training data recency using 12 months of training data at varying time intervals before the fixed test dataset.

## Results

From 01 February 2017 to 31 January 2020, 52,590 elective admissions and 202,633 emergency admissions were recorded. Using 12 pm as the prediction time, 63,909 (25.0%) short admissions were excluded from the main analyses because these admissions did not include time in hospital at 12 pm (**Figure S1**), i.e. some admissions started after 12 pm and these patients were discharged before 12 pm the following day. All other admissions <24h but spanning 12pm were included. This resulted in a total of 48,039 elective admissions (38,627 patients) and 143,275 emergency admissions (86,059 patients) included in the analyses. The median (IQR) age at admission was 65 (47-79) years, 97,869 (51.2%) patients were female, and 148,060 (77.4%) were recorded as being of white ethnicity (ethnicity missing in 33,626, 17.6%). The median (IQR) length of hospital stay was 2.2 (1.2, 5.0) days for elective admissions and 2.1 (0.8, 6.0) days for emergency admissions. The distribution of demographic characteristics was similar between the training and test datasets (**Table 2**).

**Table 2.** Baseline characteristics of 48,039 elective and 143,275 emergency admissions between 01 February 2017 to 31 January 2020 used in training and testing discharge prediction models. IMD: index of multiple deprivation (higher scores indicate greater deprivation, range 0-73.5.

|  | Elective (N= 48039) |  | Emergency (N= 143275) |  | Total (N= 191314) |
| --- | --- | --- | --- | --- | --- |
|  | Training (N= 32832) | Test (N= 15207) | Training (N= 92611) | Test (N=50664) |  |
| **Age (years)** |  |  |  |  |  |
| Median (Q1, Q3) | 62.0 (48.0, 73.0) | 62.0 (48.0, 73.0) | 67.0 (47.0, 81.0) | 66.0 (46.0, 81.0) | 65.0 (47.0, 79.0) |
| **Sex** |  |  |  |  |  |
| Female | 16409 (50.0%) | 7603 (50.0%) | 47790 (51.6%) | 26067 (51.5%) | 97869 (51.2%) |
| Male | 16423 (50.0%) | 7604 (50.0%) | 44821 (48.4%) | 24597 (48.5%) | 93445 (48.8%) |
| **Ethnicity** |  |  |  |  |  |
| White | 23977 (73.0%) | 10961 (72.1%) | 73611 (79.5%) | 39511 (78.0%) | 148060 (77.4%) |
| Mixed | 173 (0.5%) | 92 (0.6%) | 627 (0.7%) | 390 (0.8%) | 1282 (0.7%) |
| Asian | 712 (2.2%) | 402 (2.6%) | 2198 (2.4%) | 1264 (2.5%) | 4576 (2.4%) |
| Black | 355 (1.1%) | 185 (1.2%) | 1040 (1.1%) | 566 (1.1%) | 2146 (1.1%) |
| Other | 271 (0.8%) | 155 (1.0%) | 792 (0.9%) | 406 (0.8%) | 1624 (0.8%) |
| Unknown | 7344 (22.4%) | 3412 (22.4%) | 14343 (15.5%) | 8527 (16.8%) | 33626 (17.6%) |
| **IMD deprivation score** |  |  |  |  |  |
| Median (Q1, Q3) | 9.8 (5.9, 15.5) | 9.9 (6.2, 15.8) | 10.5 (6.2, 16.5) | 10.3 (6.5, 16.3) | 10.2 (6.2, 16.1) |
| Missing | 525 | 212 | 1122 | 555 | 2414 |
| **Admission source** |  |  |  |  |  |
| Usual place of residence | 32232 (98.2%) | 14959 (98.5%) | 87493 (94.6%) | 48212 (95.4%) | 182896 (95.7%) |
| Other hospital provider | 487 (1.5%) | 196 (1.3%) | 4210 (4.6%) | 2023 (4.0%) | 6916 (3.6%) |
| Other | 92 (0.3%) | 36 (0.2%) | 758 (0.8%) | 324 (0.6%) | 1210 (0.6%) |
| Missing | 21 | 16 | 150 | 105 | 292 |
| **Length of stay (days)** |  |  |  |  |  |
| Median (Q1, Q3) | 2.2 (1.2, 5.1) | 2.2 (1.2, 4.9) | 2.1 (0.9, 6.1) | 2.0 (0.8, 5.7) | 2.1 (1.0, 5.6) |

### Model performance for individual-level prediction

Predicting discharge at 12 pm, 237,672 and 809,279 patient days for elective and emergency admissions were included in the analyses. 47,177 (19.8%) and 141,531 (17.5%) discharge events within 24 hours of the index date were observed, respectively. The proportion of patients discharged from the hospital within the next 24 hours decreased as the prior length of stay in the current admission increased, and varied between emergency and elective admissions, and the day of the week of the index date (**Figure 1**).

Using the validation dataset, we evaluated model performance with varying numbers of features, ranging from 10 to all 1,152 features. Model performance initially improved as more features were included, but then plateaued with ≥200 features (and for some metrics even slightly declined) (**Figure S2**). As expected, training time increased with the number of features (**Figure S2**). We therefore used the top 200 ranked emergency and elective model features in all subsequent emergency and elective models.

For elective admissions, the AUC for predicting discharge within 24 hours was 0.871, and the AUPRC was 0.658 (**Figure S3**). Using a probability threshold that optimised F1 score in the validation dataset, the PPV was 0.555, NPV 0.911, and F1 score 0.609 (**Table 3**). The performance for emergency admissions was slightly lower than elective admissions, with an AUC of 0.860, AUPRC 0.644 (**Figure S3**), PPV 0.571, NPV 0.912, and F1 score 0.593 (**Table 3**).

**Table 3.** Model performance of the extreme gradient boosting (XGB) model and baseline logistic regression (LR) model predicting 24-hour discharge in the test dataset (01 February 2019 to 31 January 2020) and an additional test dataset post-COVID (01 February 2021 to 31 January 2022). The LR model only included age, sex, day of the week, and hours since admission as baseline comparator. PPV: positive predictive value; NPV: negative predictive value; AUC: area under the receiver operating curve; AUPRC: area under the precision-recall curve; MAE: normalised mean absolute error (mean difference in predicted and actual discharges per day divided by the mean number of discharges per day).

| Admission type | Model | Accuracy | Balanced accuracy | Sensitivity/Recall | Specificity | PPV/Precision | NPV | F1-score | AUC | AUPRC | MAE (%) |
| --- | --- | --- | --- | --- | --- | --- | --- | --- | --- | --- | --- |
| **Test data: 01/02/2019 to 31/01/2020** |  |  |  |  |  |  |  |  |  |  |  |
| Elective | XGB | 0.823 | 0.767 | 0.673 | 0.861 | 0.555 | 0.911 | 0.609 | 0.871 | 0.658 | 8.9 |
| Elective | LR | 0.464 | 0.596 | 0.820 | 0.372 | 0.252 | 0.889 | 0.385 | 0.629 | 0.269 | 10.7 |
| Emergency | XGB | 0.844 | 0.756 | 0.616 | 0.896 | 0.571 | 0.912 | 0.593 | 0.860 | 0.644 | 4.9 |
| Emergency | LR | 0.637 | 0.654 | 0.682 | 0.626 | 0.292 | 0.897 | 0.409 | 0.708 | 0.349 | 5.8 |
| Overall | XGB | 0.837 | 0.752 | 0.615 | 0.888 | 0.561 | 0.909 | 0.587 | 0.859 | 0.634 | 4.6 |
| Overall | LR | 0.589 | 0.642 | 0.726 | 0.558 | 0.276 | 0.898 | 0.400 | 0.694 | 0.327 | 5.4 |
| **Test data: 01/02/2021 to 31/01/2022** |  |  |  |  |  |  |  |  |  |  |  |
| Elective | XGB | 0.825 | 0.753 | 0.638 | 0.869 | 0.532 | 0.911 | 0.580 | 0.864 | 0.614 | 11.6 |
| Emergency | XGB | 0.835 | 0.703 | 0.501 | 0.906 | 0.528 | 0.896 | 0.514 | 0.820 | 0.543 | 10.0 |

Predicted probabilities reflected the real probabilities of discharge after calibration, with calibration errors of 0.003 and 0.001 for elective and emergency admissions, respectively (**Figure S4**). Performance in training, validation, and test data is shown in **Table S2**.

The XGB models showed substantially better performance than the baseline logistic regression model, which had AUCs of 0.629, 0.708, AUPRCs of 0.269, 0.349, and F1 scores of 0.385, 0.409 for elective and emergency admissions, respectively (**Table 3**). When combining elective and emergency admissions into a single XGB model, performance was similar to that of the XGB model for emergency admissions, with an AUC of 0.859, AUPRC 0.634, PPV 0.561, NPV 0.909, and F1 score 0.587.

When using the trained model to predict discharge in the post-COVID test data (01 February 2021 to 31 January 2022), performance remained similar for elective admissions, with an AUC of 0.864 and AUPRC of 0.614, but was lower for emergency admissions, with an AUC of 0.820 and AUPRC of 0.543 (**Table 3**).

### Model performance for hospital-level prediction

We calculated the total number of discharges expected in the next 24 hours across all elective or emergency admissions in the hospital by summing the individual-level predicted discharge probabilities. The predicted number of discharges accurately reflected daily fluctuations in discharge numbers during the week for elective and emergency patients, and the performance was similar across the whole test data period (**Figure 2**). The MAE was 8.9% (MAE=3.7 discharges, mean total discharges = 41) for elective admissions, lower than the 10.7% (MAE=4.4/41) obtained using baseline logistic regression models, and was 4.9% (MAE= 7.2/146) for emergency admissions using XGB compared with 5.8% (MAE=8.6/146) using the baseline models (**Table 3**). MAEs were higher in post-COVID test data, being 11.6% (MAE= 3.9/34) for elective (higher in percentage terms in part because of lower total discharge numbers), and 10.0% (MAE= 15.0/150) for emergency (**Table 3, Figure S5**).

**Figure 2.**
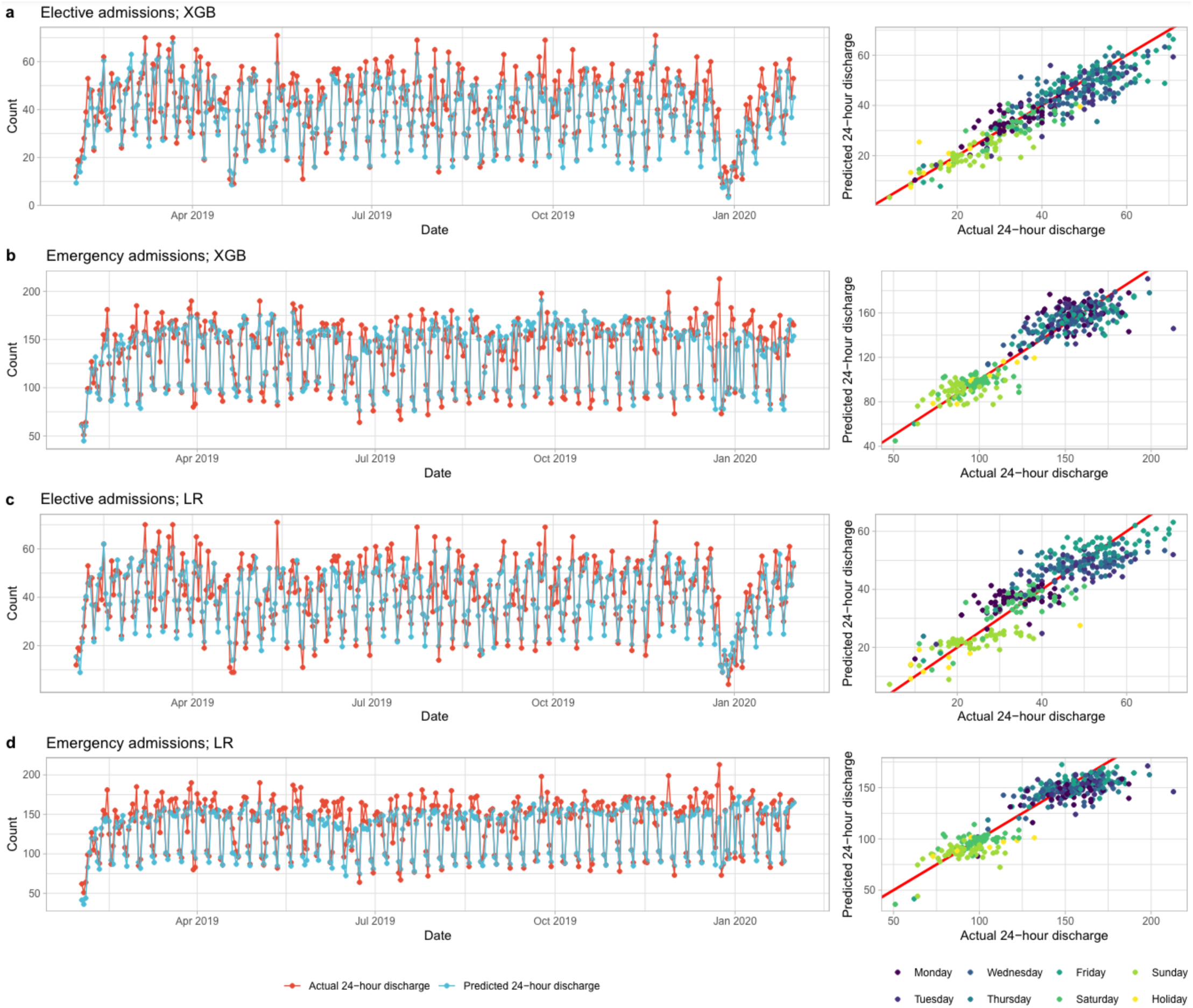
Predicted and actual number of discharges within 24 hours by calendar time in the test dataset (01 February 2019 to 31 January 2020). a) Elective admissions using extreme gradient boosting (XGB) model. b) Elective admissions using baseline logistic regression (LR) model. c) Emergency admissions using XGB model. b) Emergency admissions using baseline LR model.

### Subgroup performance and model fairness

Using balanced accuracy to jointly summarise sensitivity and specificity, model performance was broadly similar by sex, ethnicity, and deprivation score, but some variations existed in other subgroups. For elective admissions, predictions made on Monday and Sunday, and for patients admitted to trauma and orthopaedics or acute medicine had lower balanced accuracy. For emergency admissions, balanced accuracy was lower on Sundays, and for patients admitted to trauma and orthopaedics and medical subspecialties. For both elective and emergency admissions, balanced accuracy was lower in those >80 years, those with high comorbidity scores, with increasing days since admission, and in admissions from other hospital providers. Balanced accuracy was also substantially lower in those who died in hospital than those who were discharged alive (with lower AUCs, AUC= 0.712 vs 0.870, 0.762 vs 0.862 for elective and emergency admissions, respectively). In addition to lower accuracy, lower discharge rates in some subgroups including with increasing prior length of stay, also contributed to lower in PPV in those groups, albeit with linked increases in NPV (**Figure 3**, see **Figure S6** for F1 scores, AUC and AUPRC). Most subgroups met an equalised odds difference threshold of 0.1 (**Figure S6**). Within elective admissions, exceptions included predictions made on Sunday, patients admitted from other hospital providers, with ≥10 days prior length of stay, and those who died in hospital. Predictions made on the day of admission had better than average performance. For emergency admissions, exceptions included patients >80 years, those admitted to trauma and orthopaedics or from other hospital providers, with ≥4 days since admission, and those who died in hospital.

**Figure 3.**
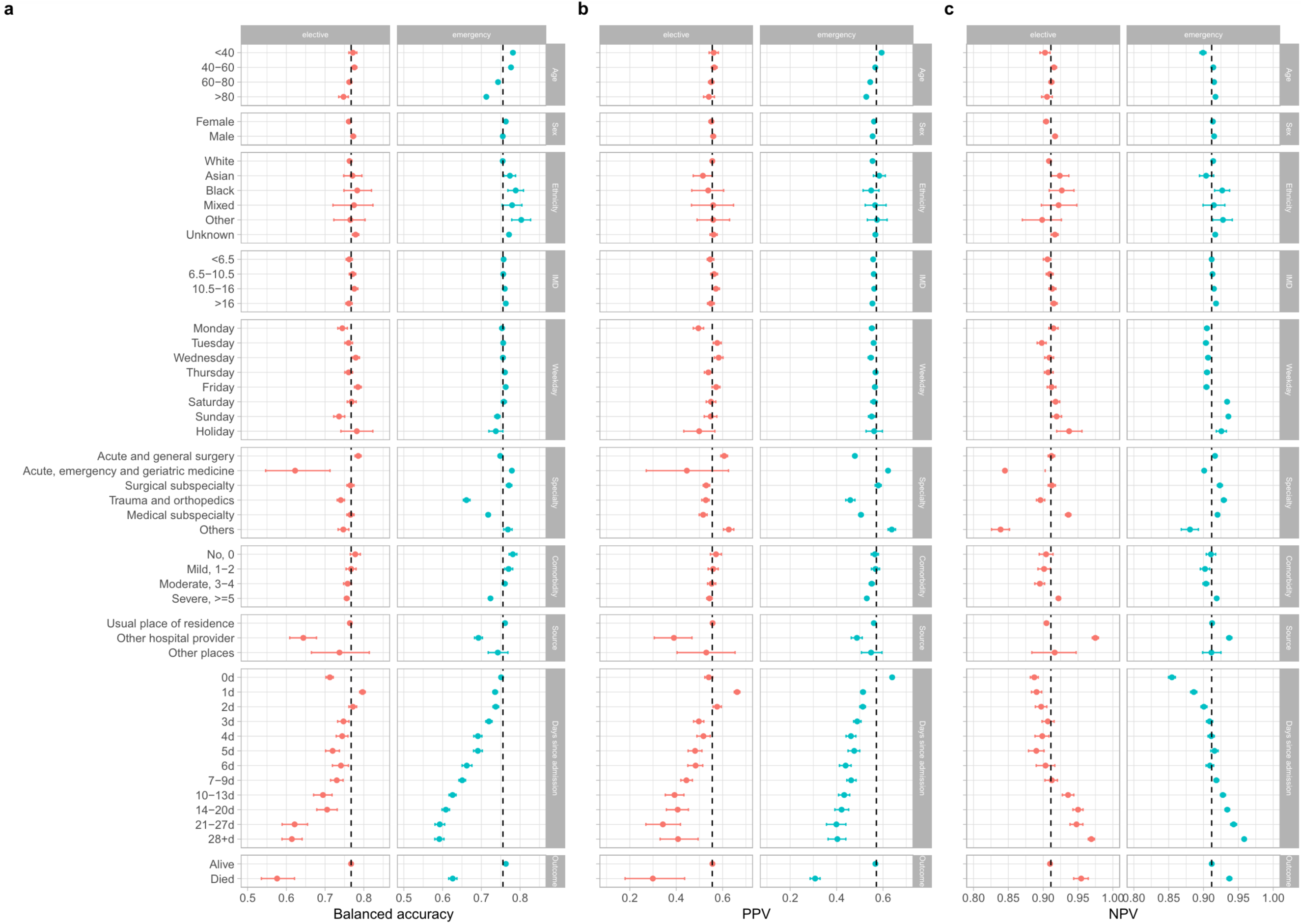
Model performance by subgroups in the test dataset (01 February 2019 to 31 January 2020). Balanced accuracy (a), positive predictive value (PPV) (b), and negative predictive value (NPV) (c) were compared. IMD=index of multiple deprivation score (higher scores indicate greater deprivation). ‘Weekday’ refers to the day of the week of the index date. Comorbidity was calculated using Charlson comorbidity score. ‘Source’ refers to the source of admission. Overall performance is shown by the dashed line in each plot. 95% confidence intervals were calculated using bootstrap. F1 score, area under the receiver operating curve (AUC), and area under the precision-recall curve (AUPRC) are shown in **Figure S6**.

### Sensitivity analyses by prediction time

Patients were more likely to be discharged between 10 am and 8 pm, and the observed proportion of patients discharged within the next 24 hours slightly varied by prediction time chosen (**Figure S7**). Using models trained at 12 pm, performance varied with different prediction times (12 am, 6 am, 12 pm, 6 pm, randomly throughout the day), with the best performance at 12 pm (AUC=0.871, 0.860 for elective and emergency admissions, respectively), followed by 6 am for elective admissions (AUC=0.861) and 6 pm for emergency admissions (AUC=0.812). Performance was lowest predicting discharges over the next 24h at 6 pm for elective admissions (AUC=0.817) and 12 am for emergency admissions (AUC=0.789) (**Figure 4**).

**Figure 4.**
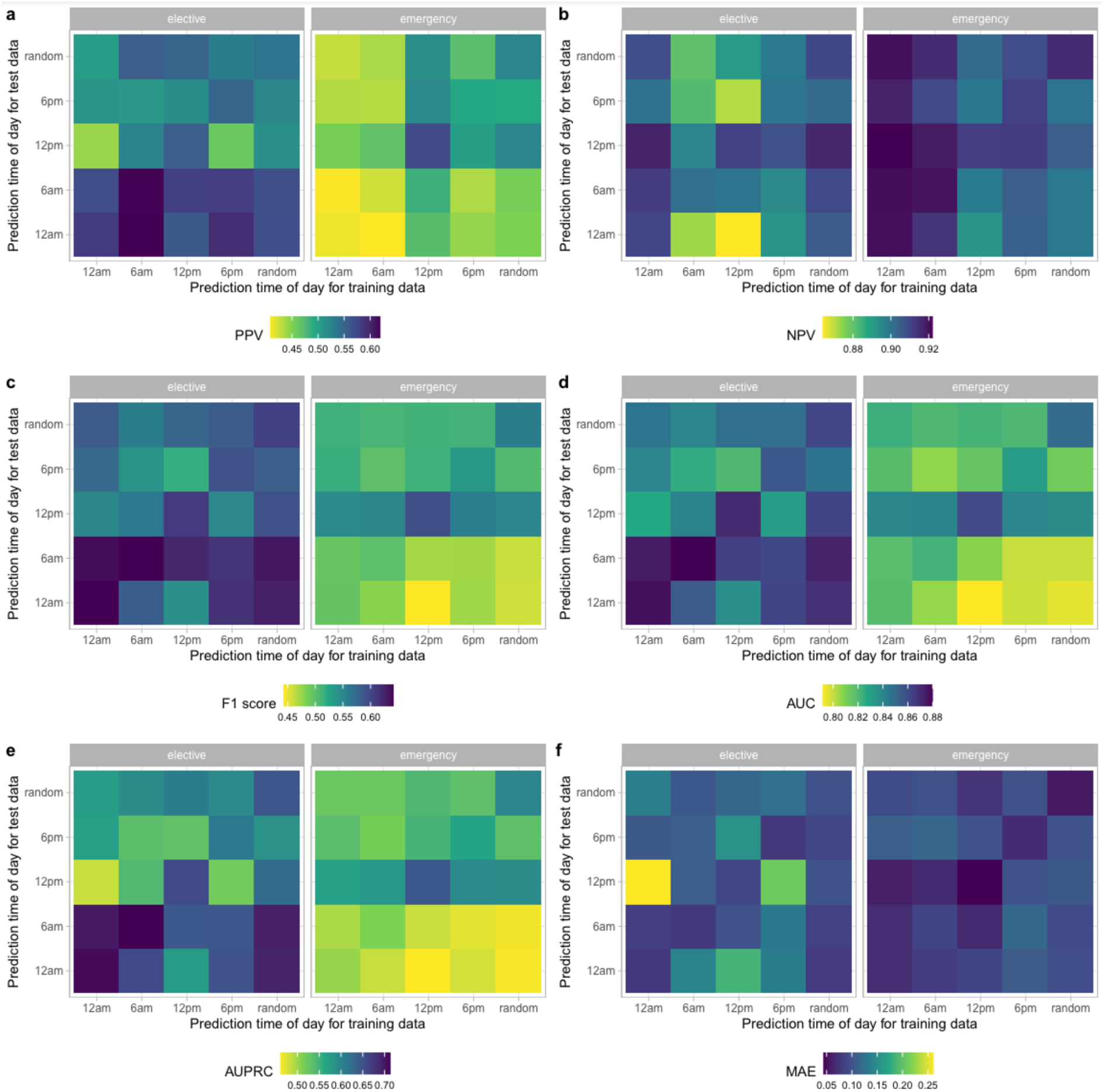
Model performance using different prediction times of day for training and test data. a) Positive predictive value. b) Negative predictive value. c) F1 score. d) Area under the receiver operating curve (AUC). e) Area under the precision-recall curve (AUPRC). f) Normalised mean absolute error (MAE) (mean difference in predicted and actual discharges per day divided by the mean number of discharges per day).

Compared to models trained and tested at 12 pm, the performance was slightly worse when the model was trained and predictions were made using times drawn randomly throughout the day, with AUC of 0.858 and 0.849 for elective and emergency admissions, respectively (**Figure 4, Figure S8**). However, this performance exceeded that observed for most models trained at a specific time but tested at different times.

When training and predicting at the same time of day, the performance was slightly better at 12 am and 6 am for elective admissions, with AUC of 0.874 and 0.876, respectively (vs. 0.871). However, the performance was lower in emergency admissions, with AUC of 0.816 and 0.819, respectively (vs. 0.860). The performance was slightly lower at 6 pm for both elective and emergency admissions compared to 12 pm (AUC=0.847 and 0.826) (**Figure 4, Figure S8).**

### Training dataset size and recency impact prediction performance

Fixing the test dataset to the 12 months from 01 February 2019, we evaluated the performance of models trained with data from the preceding 1-24 months, i.e., ranging from only 01 January 2019 to 31 January 2019 to the entire period from 01 February 2017 to 31 January 2019. Individual patient-level and hospital-wide performance all improved as the number of months of training data increased, but largely plateaued after 12 months (**Figure 5a**). This saturation effect suggested that the 24 months of training data used in the main analysis was more than sufficient to achieve optimal performance. The relative percentage change in AUC for 1 vs 24 months, and 12 vs 24 months of training data was 8% and 0.9% for elective admissions, 5% and 0.6% for emergency admissions, and in AUPRC 24% and 2.1%, and 10% and 1.6% for elective and emergency admissions, respectively.

**Figure 5.**
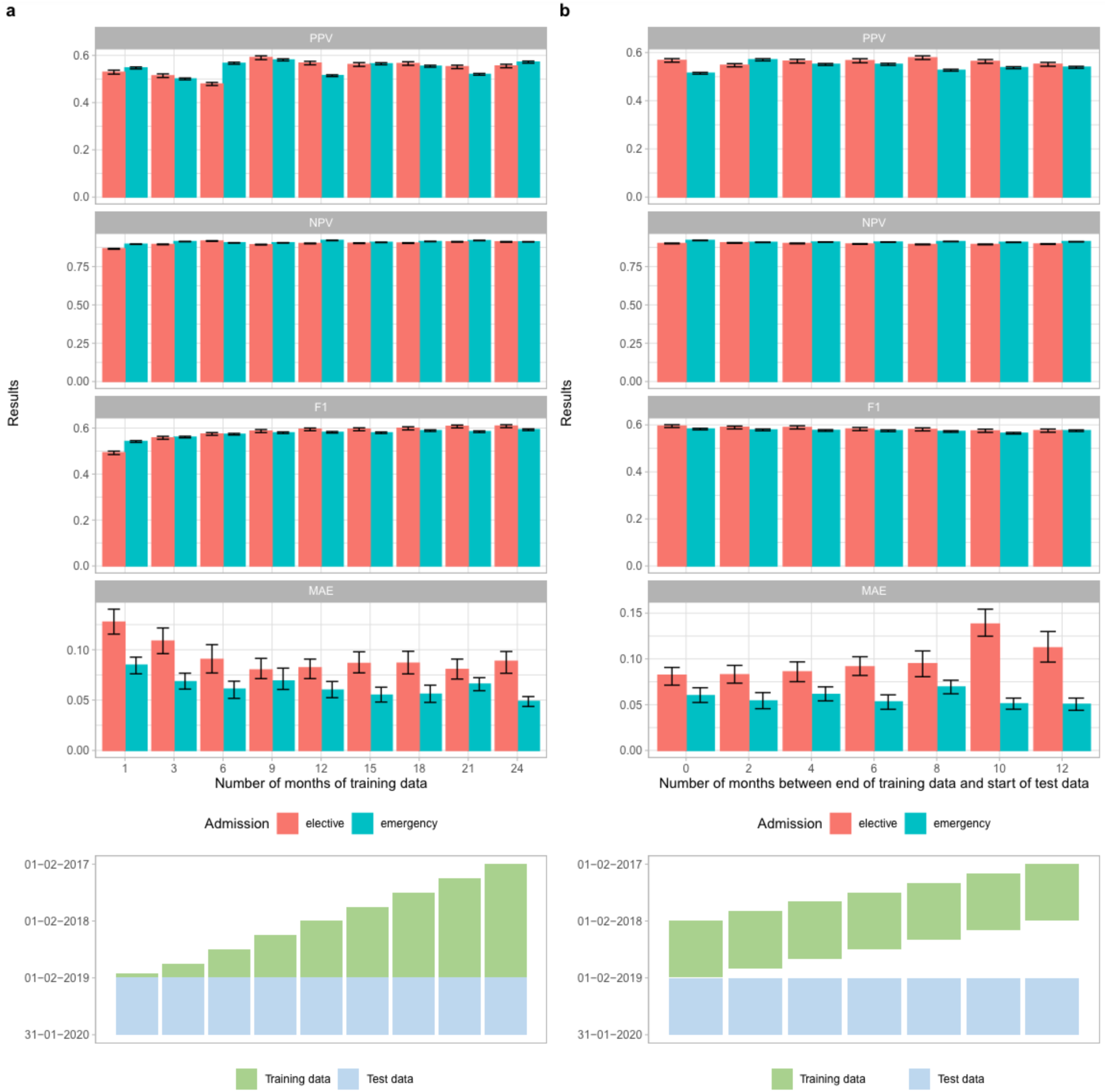
Impact of training dataset size and recency on discharge prediction performance in the test dataset (01 February 2019 to 31 January 2020). a) By increasing training data size. b) By decreasing recency of same-size training data. 95% confidence intervals were calculated using bootstrapping. PPV: positive predictive value; NPV: negative predictive value; MAE: normalised mean absolute error (mean difference in predicted and actual discharges per day divided by the mean number of discharges per day).

Mimicking real-world implementation, we also considered the impact of decreasing the recency of training data (**Figure 5b**). We used the same fixed test dataset, but only 12 months of training data varying with an interval of 0 to 12 months between the end of the training period and the start of the testing period. Performance was generally consistent, with the most recent training data performing only slightly better. The relative percentage change for 0 vs 12-month intervals in AUC was only 1% and 1%, and AUPRC was only 5% and 2% for elective and emergency admissions, respectively.

### Feature importance

The top five most important features for predicting discharge in elective admissions were number of oral medications received in the last 24 hours, the standard deviation of historic length of stay for other patients on the current ward, if the patient completed a course of antibiotics in the last 24 hours, receipt of intravenous antibiotics in the last 24 hours, and the number of procedures the patient underwent in the last 24 hours. For emergency admissions, the five most important features were number of oral medications in the last 24 hours, completion of antibiotics in the last 24 hours, hours since admission, the standard deviation of historic length of stay for other patients on the current ward, and receipt of intravenous antibiotics in the last 24 hours. The top 20 most predictive features are shown in **Figure 6**. We also grouped individual features by feature category and summarised the mean importance of the top five most important features within each feature category. The most important feature categories were (non-antibiotic) medications, antibiotics, hospital capacity factors, procedures, and lab tests in elective admissions, and (non-antibiotic) medications, antibiotics, hospital capacity factors, demographics, and current admission factors in emergency admissions (**Figure 6**). Combining elective and emergency admissions into a single model, the most important feature categories were (non-antibiotic) medications, antibiotics, hospital capacity factors, current admission factors, and ward stay features (**Figure S9**).

**Figure 6.**
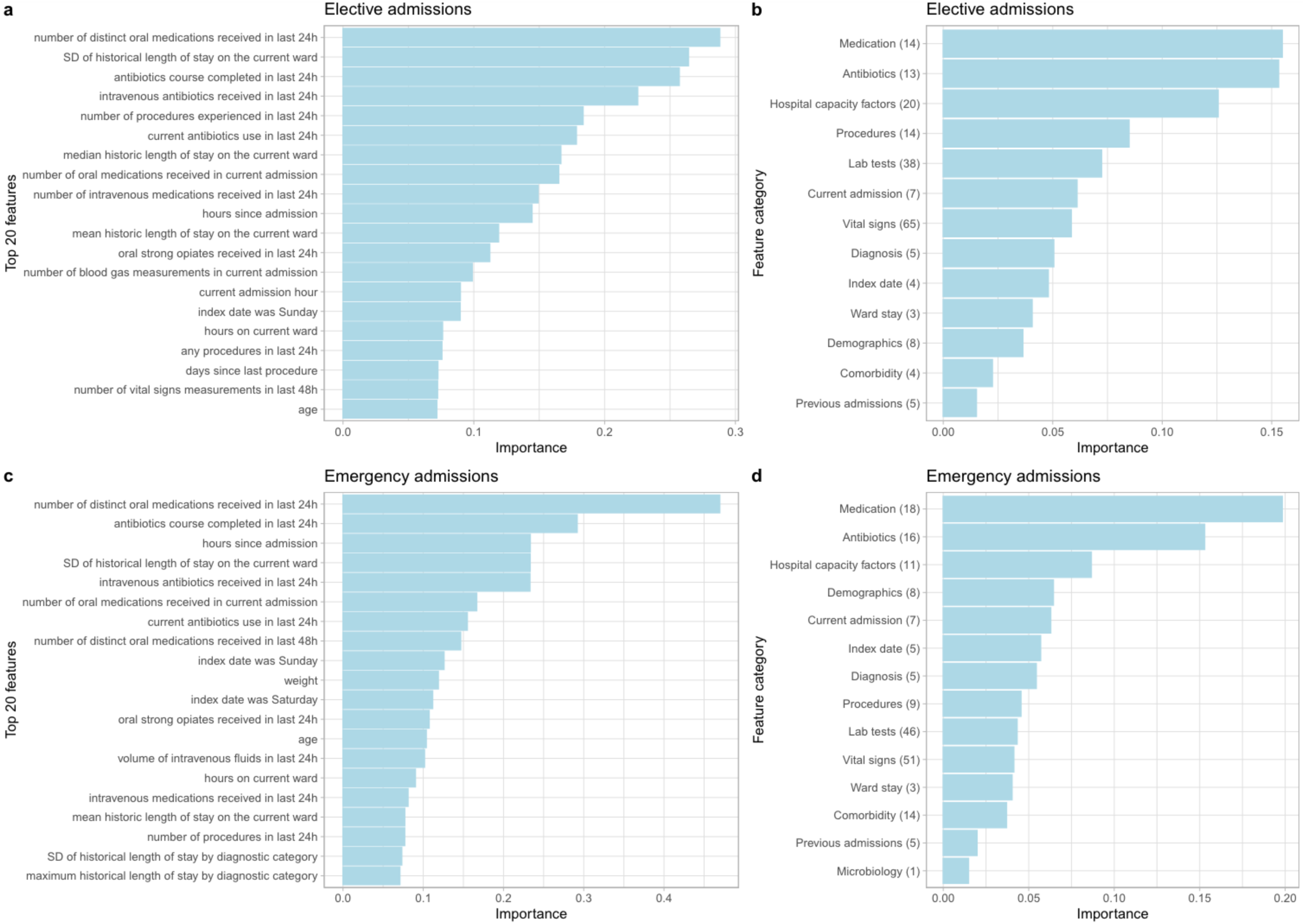
Feature importance from extreme gradient boosting models using SHAP values for elective admissions (a, b) and emergency admissions (c, d). The top 20 most predictive features are shown in the order of predictiveness in panel **a** and **c**. Feature importance grouped by feature category is shown in the order of predictiveness in panel **b** and **d.** The mean importance of the top 5 most important features within each category is plotted. Numbers shown in parenthesis are number of features within the top 200 most predictive features in each category. No microbiology features were selected for elective admissions, and no discharge planning features were selected for both admission types. The complete list of features is summarised in **Table S1.** SHAP: SHapley Additive exPlanations. SD: standard deviation; Current admission: admission time/source/specialty; diagnosis: length of stay characteristics of diagnostic categories; Previous admissions: previous length of stay and readmission; Discharge: discharge planning.

## Discussion

Machine learning underpinned by large-scale EHR data has the potential to transform how healthcare is delivered, but applications to the operational management of hospitals are largely unexplored^19^. By exploiting a wide range of features in EHR, we can accurately predict patient discharge events within the next 24 hours across hospitals. We predicted the total number of discharges each day following an elective admission with an MAE of 8.9% and 4.9% following an emergency admission. We also achieved accurate predictions for individual patients with an AUC of 0.871 and 0.858 for elective and emergency admissions, respectively. PPV and NPV were 0.555 and 0.911 following an elective admission, and 0.571 and 0.912 following an emergency admission. We achieved substantially better performance at predicting individual discharge than the baseline logistic regression model, with a 0.18-0.22 absolute improvement in F1 score, 0.15-0.24 improvement in AUC, and 0.30-0.39 improvement in AUPRC.

We built on prior studies and included a wide range of features, selected and curated by experienced clinicians, statisticians, and machine learning experts. We accounted for individual patient factors, both immediate and longer-term, and considered hospital-wide factors including historical length of stay for specific conditions. Our model performance is amongst the best reported for this task, while also pragmatically accounting for patients admitted more than once and those who died, in contrast to the best-performing reported model which excluded both these patient groups^10^. While alternative model architectures, including those that allow entire time series as inputs, might enhance performance, they may likely also need improved data quality to achieve significant performance gains at the cost of computational resources and training time. Although the model performance achieved by our study and others could translate to operational benefits, further studies comparing model performance to clinician predictions and of trial implementations are required.

We aggregated patient-level probabilities into precise predictions of daily discharge numbers at the hospital level, in contrast to other approaches modelling total discharges for the entire population^20^ or combining binary patient-level predictions (e.g., admission/discharge yes/no)^21^. Splitting data by calendar time rather than randomly splitting data into training and testing, we showed that hospital data in previous years could accurately predict discharges in future years. Also, our model was generally robust in predicting discharges following the start of the COVID-19 pandemic, with the performance for elective patients remaining essentially unchanged. However, performance was lower in emergency admissions, likely in part reflecting changes in reasons for admission, hospital capacity^22^ and the availability of community support following discharge.

Our model performance was consistent across different population subgroups by sex, ethnicity, and deprivation, but was lower in longer admissions, older patients, and those who were admitted from other hospital providers. We achieved best performance for short-stay patients, where factors related to their active treatment and response were important in discharge decisions and were relatively well captured by the data we used. In contrast, for longer-stay and older patients, particularly after an emergency admission, gaps in available data led to less accurate predictions. This highlights the importance of extending the data types available to improve performance, e.g., assessments of functional state which is often documented only as free text and were not available in the data we used, and external factors such as the availability of social care and typical waiting times for social care packages or residential care. Performance was much lower in patients who died in hospital, potentially because the model training favoured features that predicted recovery rather than deterioration when predicting discharge. Fitting a multiclass model, e.g. predicting discharge alive in 24 hours, death before discharge, and discharge alive after 24 hours, could address this, since these different outcomes are likely to have different predictors. This might substantially improve individual-level predictions whilst having less impact on overall accuracy based on summing individual-level predicted probabilities of discharge alive and death across the population.

Model performance was better with increasing training data size, with saturation at around 12 months’ training data, and was slightly better using more recent than distant data, suggesting that training could be undertaken without using excessive historical data and could simply be updated two or three times a year in a real-world application. Additionally, differences between the time of day that models were trained and tested on had a relatively modest impact, however where computational resources allow, optimal performance could be achieved by using models tuned to specific times of day.

We found that non-antibiotic medications, antibiotics, and hospital capacity factors were the most predictive feature categories for both emergency and elective admissions. Switching to oral medications or completing antibiotic courses usually indicates clinical improvement, and hospital capacity factors such as the length of stay of current inpatients and the number of patients in the current ward reflect the crowdedness/pressure on hospital beds. On the contrary, discharge planning (physiotherapy contacts), microbiology tests, and previous lengths of stay and readmissions played a relatively minor role in discharge prediction.

Sequentially including more of the most predictive features increased model performance, but plateaued after including the top 200 features, and the computational time was substantially reduced to only 20% vs when all 1152 features were used, suggesting that including more features does not necessarily improve performance. Hospitals may need to pay more attention to the data collection quality of the key features that are most predictive to achieve accurate and efficient predictions.

Ensuring the availability of beds and timely patient discharge is pivotal in managing patient flow within healthcare systems^23^, but existing interventions often adopt static procedures such as ‘discharge by noon’, or respond only to critical levels of patient demand^24^, therefore failing to address the broader complexities of patient flow dynamics. Moreover, the integration of automated discharge prediction models into clinical practice remains limited^25^. We envision our approach could be deployed in several ways. For example, within each hospital ward, a dashboard of discharge tasks (follow up plan, discharge letters, discharge medication, transport, social care readiness, etc) and binary predictions of discharge within the next 24 hours, could be used by healthcare workers to flag patients likely to be discharged but with outstanding tasks. Discharge probabilities could also be used to rank the patients most likely to go home to ensure their discharge preparations were prioritised.

Where the total number of planned or expected admissions within the next 24 hours exceeded the predicted number of discharges, preparations for redeploying staff and resources could be made by operational teams. On-going oversight of input data quality/completeness and the accuracy of predictions by specialist data science teams would be required at a hospital level. Taken together, our models have the potential to be applied to improve the efficiency of patient flow across hospital settings, leading to accelerated care delivery and patient recovery, and optimal use of healthcare resources. Although our study was based on data from hospitals in Oxfordshire, this framework could potentially also apply to hospitals in other regions within the UK and internationally with similar settings, offering a prospect for mitigating the pressure of overcrowding on NHS and other healthcare systems, thus improving healthcare delivery efficacy and resource management on a national scale.

Limitations of our study include our focus on a relatively short prediction horizon of 24 hours, limiting the scope of planning and interventions that could follow the same timescale. However, our approach could be adapted to make longer-range predictions too. We used diagnosis categories derived from ICD-10 codes for training and testing, which were only recorded at discharge, however in reality the primary working diagnosis is known in real-time to clinicians and could be used if documented electronically. We did not incorporate hospital features such as the percentage of occupied beds, as the number of available beds in each hospital was not available in our dataset. Additionally, we did not evaluate how model performance varied with the degree of operational pressure experienced by hospitals, which could be considered in the future as accurate predictions are particularly beneficial when resources are most constrained. We adopted a relatively simple, pragmatic approach to feature engineering, using summaries of time series data including blood tests and vital sign measurements. Performance could potentially be improved by better representing these dynamic data in future work, and by investigating the benefits of dimensionality reduction and more complex feature representations such as *t*-SNE^26^ or autoencoders^27^. Moreover, we used temporal data from the same hospital for validation and testing, rather than data from a completely different hospital group. Future work should incorporate validation with data from diverse settings to further strengthen the validity and generalisability of our findings, as well as studies of the impact of deploying similar models.

We only used structured EHR data for predicting imminent hospital discharge and did not consider other data types such as unstructured free text, which could potentially improve prediction further, particularly for patients with prolonged hospital stays. We used the XGBoost algorithm, which is a widely employed method recognised for its superior performance compared to other traditional machine learning models, but other advanced architectures including deep learning models have the potential for improving performance and accommodating flexible updates, such as incorporating new data over time or across different settings. In recent years, there has been an increasing interest in using natural language processing and deep learning models for hospital management tasks such as length of stay prediction and utilising more complex data including free text medical records^28–30^. For example, a recent study demonstrated the efficacy of large language models trained on unstructured clinical notes for predicting hospital length of stay, outperforming traditional models^31^. Future studies should explore similar approaches and make use of unstructured data to enhance predictive capabilities for healthcare management.

In conclusion, our study shows the feasibility of integrating machine learning modelling approaches with EHR data to facilitate real-time operational management in hospitals, with realistic requirements for training data and model updating. Our models achieve a good performance for both individual-level and hospital-level discharge predictions, demonstrating the potential to be deployed to improve the efficiency of hospital management, patient flow dynamics, and expedite patients’ recovery and discharge processes.

## Data availability

The datasets analysed during the current study are not publicly available as they contain personal data but are available from the Infections in Oxfordshire Research Database (https://oxfordbrc.nihr.ac.uk/research-themes/modernising-medical-microbiology-and-big-infection-diagnostics/iord-about/), subject to an application and research proposal meeting the ethical and governance requirements of the Database. For further details on how to apply for access to the data and a research proposal template please.

## Code availability

A copy of the analysis code is available at https://github.com/jiaweioxford/discharge_prediction.

## Ethics Committee Approval

Deidentified data were obtained from the Infections in Oxfordshire Research Database which has approvals from the National Research Ethics Service South Central – Oxford C Research Ethics Committee (19/SC/0403), the Health Research Authority and the National Confidentiality Advisory Group (19/CAG/0144), including provision for use of pseudonymised routinely collected data without individual patient consent. Patients who choose to opt out of their data being used in research are not included in the study. The study was carried out in accordance with all relevant guidelines and regulations.

## Acknowledgements

This work uses data provided by patients and collected by the UK’s National Health Service as part of their care and support. We thank all the people of Oxfordshire who contribute to the Infections in Oxfordshire Research Database. Research Database Team: L Butcher, H Boseley, C Crichton, DW Crook, DW Eyre, O Freeman, J Gearing (community), R Harrington, K Jeffery, M Landray, A Pal, TEA Peto, TP Quan, J Robinson (community), J Sellors, B Shine, AS Walker, D Waller. Patient and Public Panel: M Ahmed, G Blower, J Hopkins, V Lekkos, R Mandunya, S Markham, B Nichols. Members of the panel contribute to setting research priorities and questions, and have reviewed a summary of the data used and of the analytical approach. All results are shared with the panel for comment. We would also like to thank Lisa Glynn and colleagues at Oxfordshire University Hospitals NHS Trust for helpful feedback and insights into early versions of the data analyses presented.

## Author Contributions

The study was designed and conceived by DWE, ASW, DAC, and AJB. JW, DWE, and JZ curated the data. JW, JZ, ZZ, KY, QG, and AL analysed the data and created the visualisations. JW, JZ, and DWE wrote the first draft of the manuscript. All authors contributed to editing and revising the manuscript.

## References

1. Kruk, M. E. et al. High-quality health systems in the Sustainable Development Goals era: time for a revolution. Lancet Glob. Health 6, e1196–e1252 (2018).

2. Providers Deliver: Patient flow. https://nhsproviders.org/providers-deliver-patient-flow/introduction.

3. Nine steps to improving patient flow in UK hospitals | Health Business. https://healthbusinessuk.net/features/nine-steps-improving-patient-flow-uk-hospitals.

4. Arsenault, C. et al. COVID-19 and resilience of healthcare systems in ten countries. Nat. Med. 28, 1314–1324 (2022).

5. 5. NEJM Catalyst. What Is Patient Flow? Catal. Carryover 4, (2018).

6. Haraden, C. & Resar, R. Patient Flow in Hospitals: Understanding and Controlling It Better. Front. Health Serv. Manage. 20, 3–15 (2004).

7. Safavi, K. C. et al. Development and Validation of a Machine Learning Model to Aid Discharge Processes for Inpatient Surgical Care. *JAMA Netw*. Open 2, e1917221 (2019).

8. Lazar, D. J., Kia, A., Freeman, R. & Divino, C. M. A Machine Learning Model Enhances Prediction of Discharge for Surgical Patients. J. Am. Coll. Surg. 231, S132 (2020).

9. Ahn, I. et al. Machine Learning–Based Hospital Discharge Prediction for Patients With Cardiovascular Diseases: Development and Usability Study. JMIR Med. Inform. 9, e32662 (2021).

10. Zhang, X., Yan, C., Malin, B. A., Patel, M. B. & Chen, Y. Predicting next-day discharge via electronic health record access logs. J. Am. Med. Inform. Assoc. 28, 2670–2680 (2021).

11. Barnes, S., Hamrock, E., Toerper, M., Siddiqui, S. & Levin, S. Real-time prediction of inpatient length of stay for discharge prioritization. J. Am. Med. Inform. Assoc. 23, e2–e10 (2016).

12. Levin, S. et al. Machine-learning-based hospital discharge predictions can support multidisciplinary rounds and decrease hospital length-of-stay. BMJ Innov. 7, (2021).

13. Bertsimas, D., Pauphilet, J., Stevens, J. & Tandon, M. Predicting Inpatient Flow at a Major Hospital Using Interpretable Analytics. Manuf. Serv. Oper. Manag. 24, 2809–2824 (2022).

14. Ward, A. et al. Operationally-Informed Hospital-Wide Discharge Prediction Using Machine Learning. in 2020 *IEEE International Conference on E-health Networking*, Application & Services (HEALTHCOM) 1–6 (2021). doi:10.1109/HEALTHCOM49281.2021.9399025.

15. Niculescu-Mizil, A. & Caruana, R. A. Obtaining Calibrated Probabilities from Boosting. Preprint at 10.48550/arXiv.1207.1403 (2012).

16. Niculescu-Mizil, A. & Caruana, R. Predicting good probabilities with supervised learning. in Proceedings of the 22nd international conference on Machine learning 625–632 (Association for Computing Machinery, New York, NY, USA, 2005). doi:10.1145/1102351.1102430.

17. Lundberg, S. M. & Lee, S.-I. A Unified Approach to Interpreting Model Predictions. In Advances in Neural Information Processing Systems vol. 30 (Curran Associates, Inc., 2017).

18. Bolton, W. J. et al. Personalising intravenous to oral antibiotic switch decision making through fair interpretable machine learning. Nat. Commun. 15, 506 (2024).

19. Pianykh, O. S. et al. Improving healthcare operations management with machine learning. *Nat*. Mach. Intell. 2, 266–273 (2020).

20. van Walraven, C. & Forster, A. J. The TEND (Tomorrow’s Expected Number of Discharges) Model Accurately Predicted the Number of Patients Who Were Discharged from the Hospital the Next Day. J. Hosp. Med. 13, 158–163 (2018).

21. Sun, Y., Heng, B. H., Tay, S. Y. & Seow, E. Predicting hospital admissions at emergency department triage using routine administrative data. Acad. Emerg. Med. Off. J. Soc. Acad. Emerg. Med. 18, 844–850 (2011).

22. McCabe, R. et al. Adapting hospital capacity to meet changing demands during the COVID-19 pandemic. BMC Med. 18, 329 (2020).

23. Hoot, N. R. & Aronsky, D. Systematic review of emergency department crowding: causes, effects, and solutions. Ann. Emerg. Med. 52, 126–136 (2008).

24. Viccellio, A., Santora, C., Singer, A. J., Thode, H. C. & Henry, M. C. The association between transfer of emergency department boarders to inpatient hallways and mortality: a 4-year experience. Ann. Emerg. Med. 54, 487–491 (2009).

25. Seneviratne, M. G., Shah, N. H. & Chu, L. Bridging the implementation gap of machine learning in healthcare. BMJ Innov. 6, (2020).

26. Maaten, L. van der & Hinton, G. Visualizing Data using t-SNE. J. Mach. Learn. Res. 9, 2579–2605 (2008).

27. Zhang, L., Qi, G.-J., Wang, L. & Luo, J. AET vs. AED: Unsupervised Representation Learning by Auto-Encoding Transformations rather than Data. Preprint at 10.48550/arXiv.1901.04596 (2019).

28. Bacchi, S. et al. Prediction of general medical admission length of stay with natural language processing and deep learning: a pilot study. Intern. Emerg. Med. 15, 989–995 (2020).

29. Chen, J. et al. Multi-modal learning for inpatient length of stay prediction. Comput. Biol. Med. 171, 108121 (2024).

30. Chen, J., Qi, T. D., Vu, J. & Wen, Y. A deep learning approach for inpatient length of stay and mortality prediction. J. Biomed. Inform. 147, 104526 (2023).

31. Jiang, L. Y. et al. Health system-scale language models are all-purpose prediction engines. Nature 619, 357–362 (2023).

