## Supplementary material for "Improving patient flow through hospitals with machine learning based discharge prediction"

### Supplementary methods

#### Feature selection and data pre-processing

A total of 1,152 features were created and grouped into 15 feature categories (**Table S1),** including index date-related features, demographics, comorbidities, current admission, ward stay, current diagnostic category, procedures, antibiotics prescriptions, medication, microbiology tests, radiology investigation, readmissions and previous hospital stay, hospital capacity factors reflecting crowdedness, vital signs, and laboratory tests.

We used the Summary Hospital-level Mortality Indicator (SHMI) diagnosis group of primary diagnostic ICD10 codes[1] to summarise the expected length of stay (LOS) by diagnostic category, using data from all hospital admissions within the training dataset (01 February 2017 to 31 January 2019). We included as features the mean, standard deviation, median, maximum, and minimum of the LOS for each diagnostic category, to capture the effects of a current diagnostic category on future discharge probability[2]. We only used the training data to calculate the LOS characteristics of each diagnosis category, even when applying these estimates to the test dataset, to avoid possible data leakage, i.e., avoiding revealing information to the model that gives it an unrealistic advantage to make better predictions. ICD10 codes were assigned at discharge but were used as a proxy for the clinician's working diagnosis (not available in our dataset) to inform model predictions in real time.
For vital signs and laboratory tests, we used both numerical values reflecting the measurements themselves and the number of measurements within a particular time window, reflecting the fact that the decision to measure a vital sign or laboratory test is potentially informative in addition to the actual result obtained[3]. For example, clinicians may order additional laboratory measurements or record vital signs more frequently if patients are unstable[4]. To reduce collinearity, we grouped the number of measurements for vital signs (heart rate, respiratory rate, systolic blood pressure, diastolic blood pressure, temperature, oxygen saturation, O2 L/min, O2 delivery device, AVPU score, NEWS2 score), full blood counts (haemoglobin, haematocrit, mean cell volume, white cell count, platelets, neutrophils, lymphocytes, eosinophils, monocytes, basophils), renal function (creatinine, urea, potassium, sodium, estimated glomerular filtration rate (eGFR)), liver function (alkaline phosphatase, aspartate aminotransferase, alanine transaminase, albumin, bilirubin), bone profiles (adjusted calcium, magnesium, phosphate), clotting (activated partial thromboplastin time, prothrombin time), blood gases (base excess, partial pressure of oxygen, partial pressure of carbon dioxide, lactate, arterial blood pH), and lipids (triglycerides, high-density lipoprotein cholesterol, total cholesterol, low-density lipoprotein cholesterol), respectively. The number of measurements for other blood tests were included individually.

We pre-processed the data by setting implausible extreme values not compatible with life to missing, e.g. height 10m, temperature 20°C. Categorical features were one-hot encoded. We did not perform data truncation and standardisation as decision trees-based algorithms are insensitive to the scale of the features, and we did not perform imputation of missing values because extreme gradient boosting (XGB) models can handle missing values by default[5]. For the baseline logistic regression model, the included features (age, sex, day of the week, and hours since admission) did not have missing values.

### Prediction model

We used XGB models to predict hospital discharge within the next 24 hours. Gradient-boosted trees are an additive method iteratively combining fitted decision trees with weak individual predictive performance, into a single high-performance ensemble model after proportionally weighting individual tree contributions[6]. Each fitted decision tree (base learner with low complexity) targets the prediction residuals of the preceding tree. That is, at each step a new decision tree is built to predict the residuals, i.e., the fraction of the output not well explained by the current tree model. In this way, a new tree model is continuously added to the current collection to correct the mistakes made by the previous one. Such a sequential training approach in gradient-boosted trees is different from the independent training approach in random forests. The learning rate (between zero and one) is the most important hyperparameter in gradient-boosted trees, which controls the contribution of a new tree to the overall model. We used 5-fold cross validation within the training data and Bayesian hyperparameter optimization to select optimal model hyperparameters, including learning rate (learning_rate), the number of trees (n_estimators), the fraction of features to use (colsample_bytree), the fraction of observations to subsample at each step (subsample), maximum number of nodes allowed from the root to the farthest leaf of a tree (max_depth), minimum weight required to create a new node in a tree (min_child_weight), the minimum loss reduction required to make a split (gamma), L1 regularisation term on weight (reg_alpha), and L2 regularisation term on weight (reg_lambda). Unlike grid search and random search, which independently tune hyperparameters, Bayesian hyperparameter optimization is an informed search algorithm that each iteration learns from previous iterations and combines with the prior distribution to update the posterior of the optimization function[7]. Hyperparameter optimisation was undertaken separately for each model fitted. The final hyperparameter choices for the main models are shown in **Table S3**.

### Software

Data processing and analyses were performed in Python 3.11 using the following packages: numpy (version 1.26.4), pandas (version 2.2.0), scipy (version 1.12.0), scikit-learn (version 1.4.1), xgboost (version 2.0.3), hyperopt (version 0.2.7), and in R (version 4.3.2) using the following packages: cowplot (version 1.1.1), timeDate (version 4021.104), ggplot2 (version 3.4.4), and tidyverse (version 1.3.2).

### References

1. About the Summary Hospital-level Mortality Indicator (SHMI). *NHS Digital* https://digital.nhs.uk/data-and-information/publications/ci-hub/summary-hospital-level-mortality-indicator-shmi.
2. Bishop, J. A. *et al.* Improving patient flow during infectious disease outbreaks using machine learning for real-time prediction of patient readiness for discharge. *PLoS ONE* **16**, e0260476 (2021).
3. Biases in electronic health record data due to processes within the healthcare system: retrospective observational study | The BMJ. https://www.bmj.com/content/361/bmj.k1479.
4. Sauer, C. M. *et al.* Leveraging electronic health records for data science: common pitfalls and how to avoid them. *Lancet Digit. Health* **4**, e893–e898 (2022).
5. XGBoost Documentation — xgboost 2.0.3 documentation. https://xgboost.readthedocs.io/en/stable/index.html.
6. Hastie, T., Tibshirani, R. & Friedman, J. Boosting and Additive Trees. in *The Elements of Statistical Learning: Data Mining, Inference, and Prediction* (eds. Hastie, T., Tibshirani, R. & Friedman, J.) 337–387 (Springer, New York, NY, 2009). doi:10.1007/978-0-387-84858-7_10.
7. Wu, J. *et al.* Hyperparameter Optimization for Machine Learning Models Based on Bayesian Optimizationb. *J. Electron. Sci. Technol.* **17**, 26–40 (2019).
8. Charlson, M., Szatrowski, T. P., Peterson, J. & Gold, J. Validation of a combined comorbidity index. *J. Clin. Epidemiol.* **47**, 1245–1251 (1994).

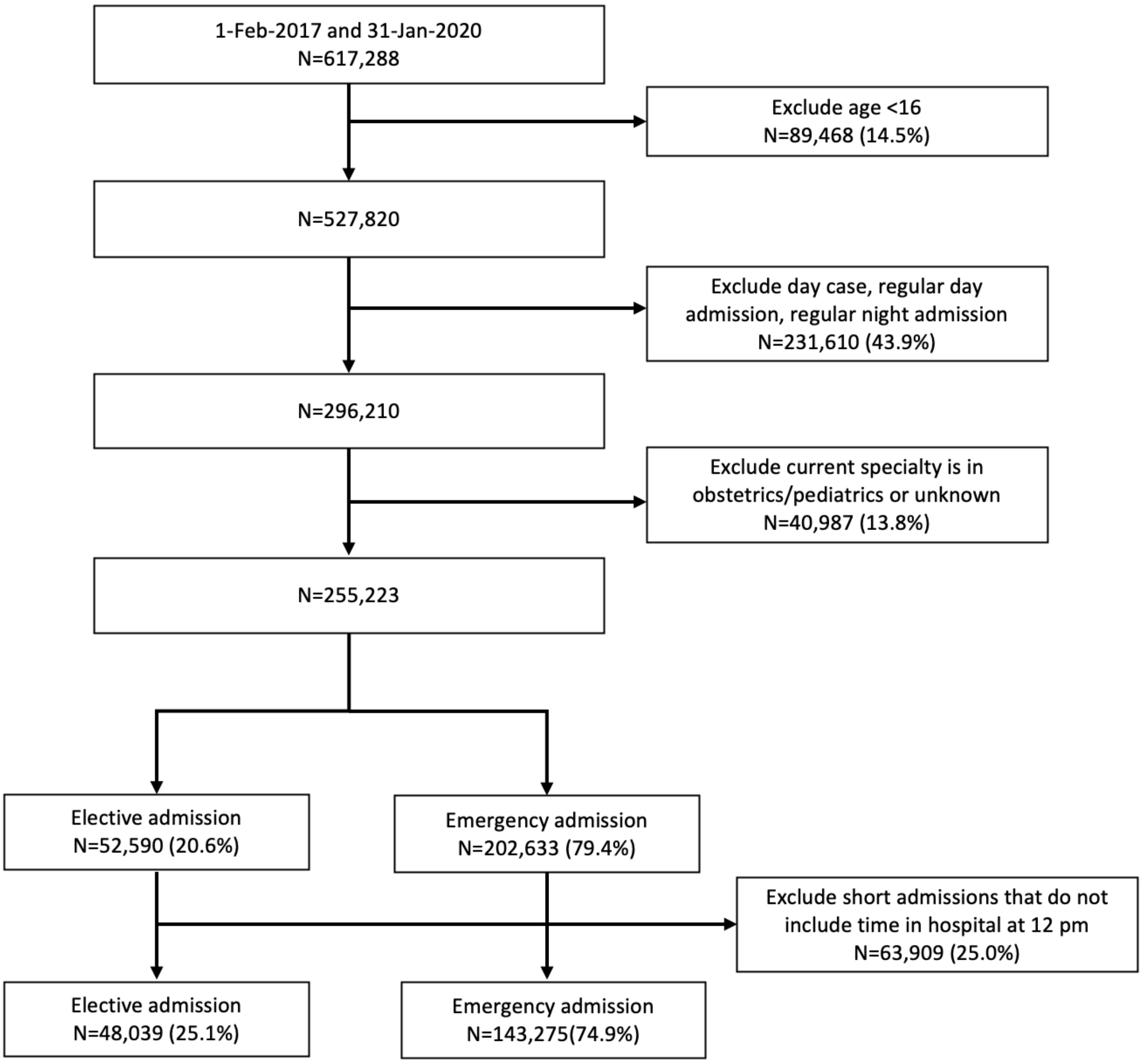

**Figure S1. Study inclusion and exclusion flow chart.**

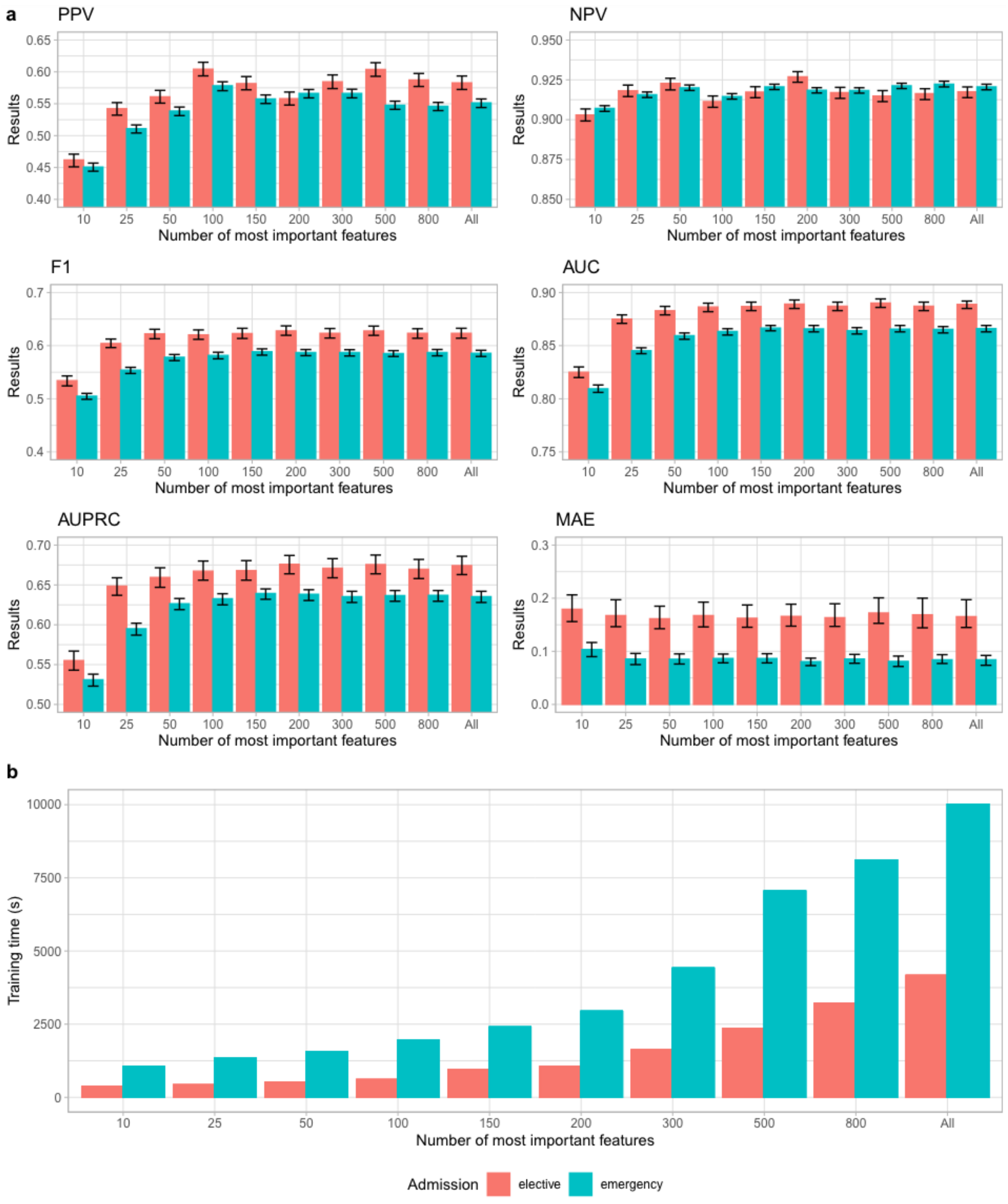

**Figure S2. Impact of number of features on model performance (a) and training time (b).** The top 10, 25, 50, 100, 150, 200, 300, and 500 most important features from the main models predicting elective and emergency discharges were selected and used for model training, respectively. 'All' included 1,152 features in total. AUC: area under the receiver operating curve; AUPRC: area under the precision-recall curve; MAE: normalised mean absolute error (mean difference in predicted and actual discharges per day divided by the mean number of discharges per day).

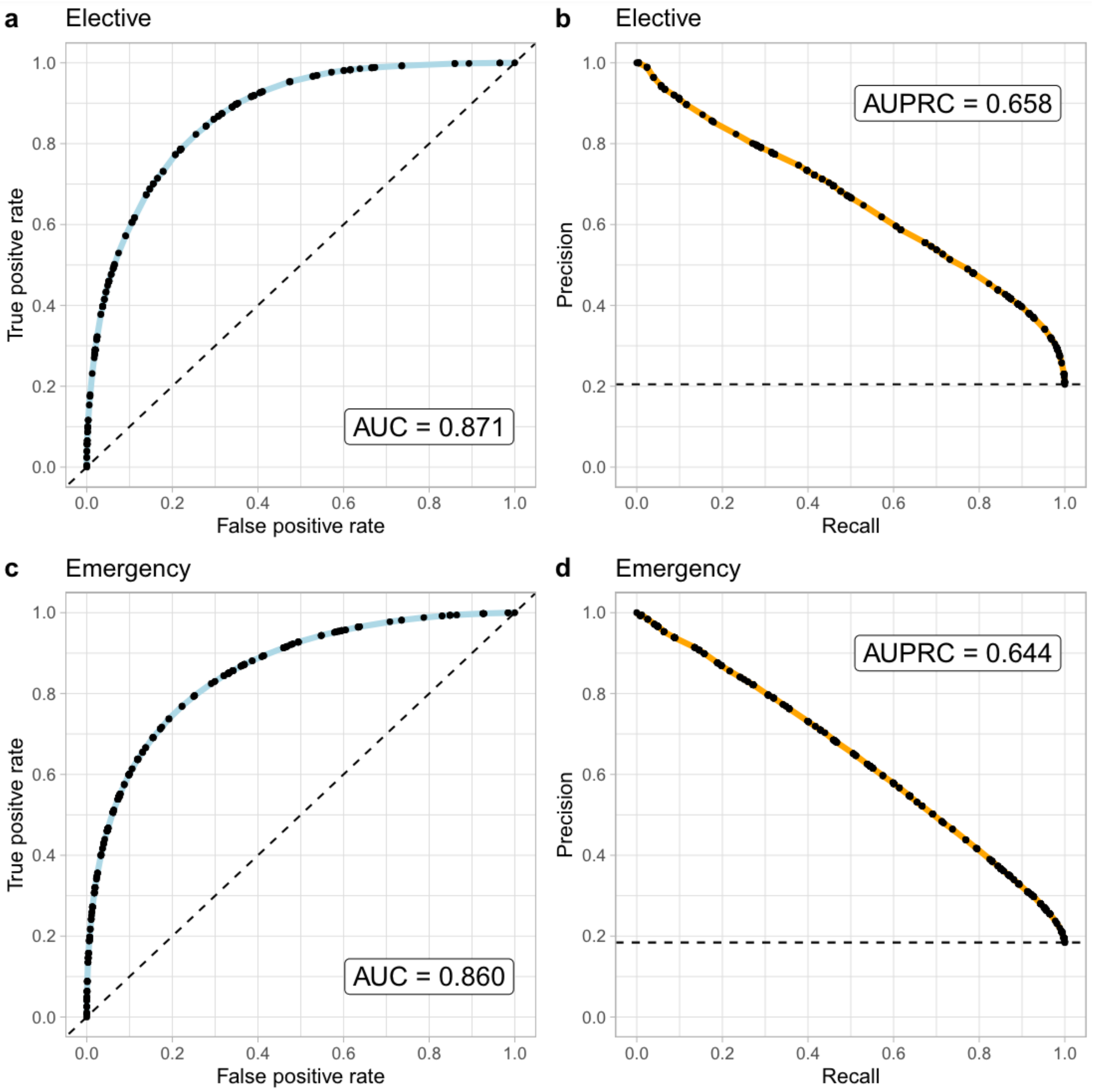

**Figure S3. Model performance of the extreme gradient boosting models in the test dataset (01 February 2019 to 31 January 2020).** a) Area under the receiver operating curve (AUC) for elective admissions. b) Area under the precision-recall curve (AUPRC) for elective admissions. c) AUC for emergency admissions. d) AUPRC for emergency admissions.

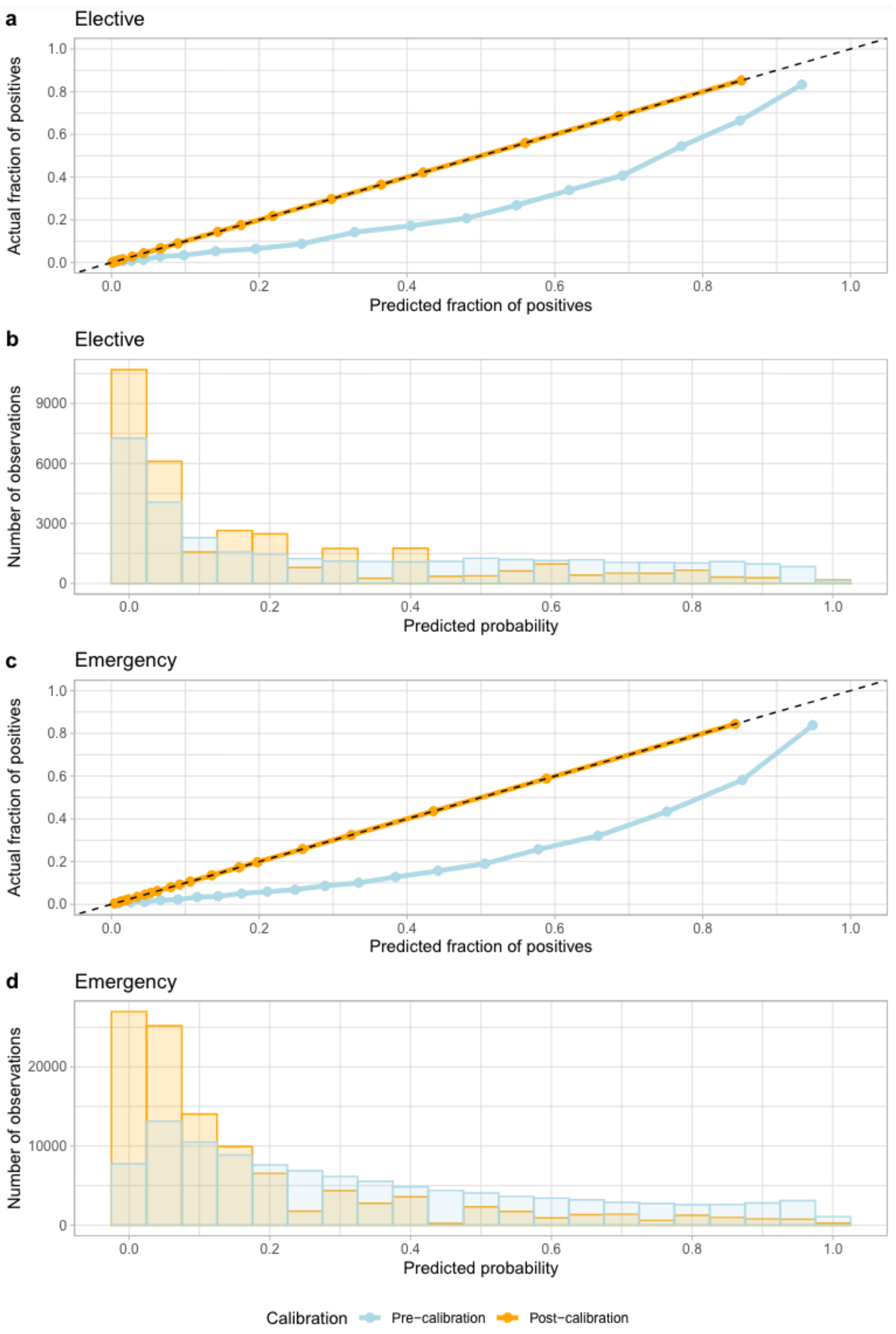

**Figure S4. Calibration curve for the extreme gradient boosting models using the validation dataset for elective admissions (a) and emergency admissions (c), and distribution of predicted probabilities pre- and post-calibration for elective admissions (b) and emergency admissions (d).** The calibration error was 0.152 and 0.003 pre/post-calibration for elective admission, and 0.203 and 0.001 pre/post-calibration for emergency admission, respectively.

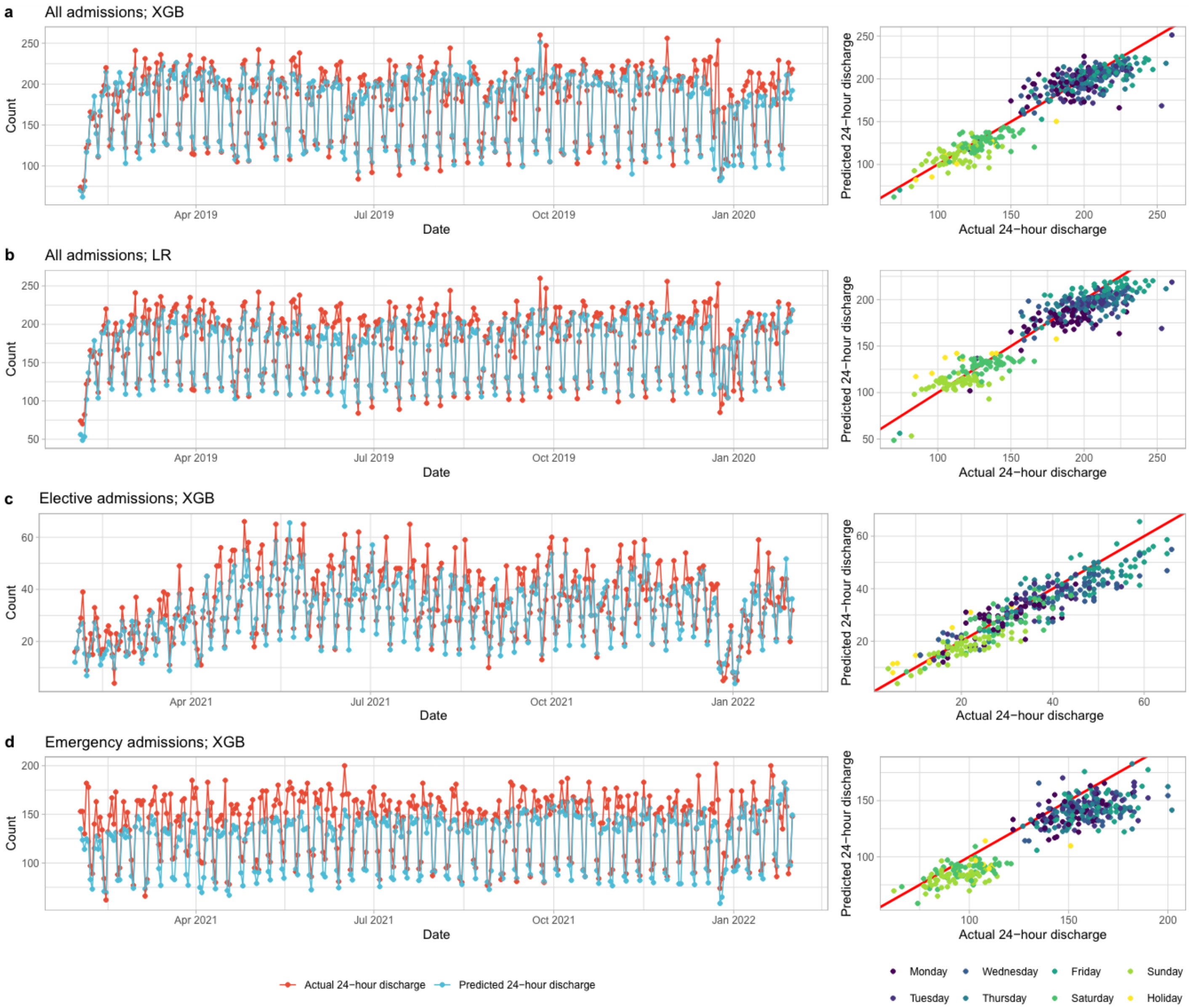

a
All admissions; XGB
b
All admissions; LR
c
Elective admissions; XGB
d
Emergency admissions; XGB
Count
Date
Apr 2019
Jul 2019
Oct 2019
Jan 2020
Apr 2021
Jul 2021
Oct 2021
Jan 2022
Predicted 24−hour discharge
Actual 24−hour discharge
Actual 24−hour discharge
Predicted 24−hour discharge
Monday
Tuesday
Wednesday
Thursday
Friday
Saturday
Sunday
Holiday

**Figure S5. Sensitivity analyses, predicted and actual number of discharges within 24 hours by calendar time in the test dataset**. a) All admissions (elective and emergency) using a single extreme gradient boosting (XGB) model in the test dataset (01 February 2019 to 31 January 2020)**.** b) All admissions using baseline logistic regression (LR) model in the test dataset (01 February 2019 to 31 January 2020). c) Elective admissions using XGB model in the post-COVID test dataset (01 February 2021 to 31 January 2022). d) Emergency admissions using XGB model in the post-COVID test dataset (01 February 2021 to 31 January 2022).

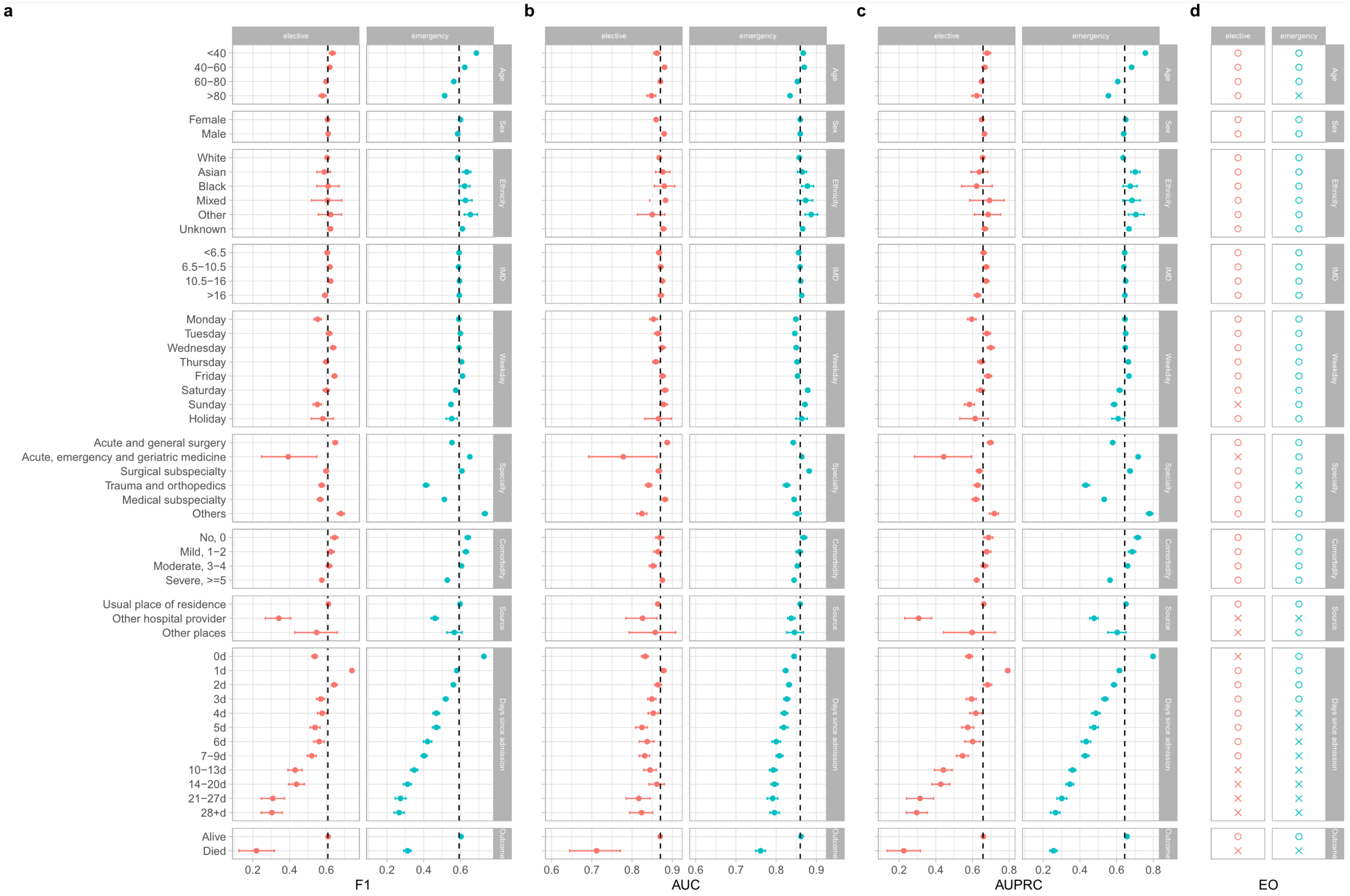
a
b
c
d
elective
emergency
Age
Sex
Ethnicity
IMD
Weekday
Specialty
Comorbidity
Source
Days since admission
Outcome
<40
40−60
60−80
>80
Female
Male
White
Asian
Black
Mixed
Other
Unknown
<6.5
6.5−10.5
10.5−16
>16
Monday
Tuesday
Wednesday
Thursday
Friday
Saturday
Sunday
Holiday
Acute and general surgery
Acute, emergency and geriatric medicine
Surgical subspecialty
Trauma and orthopedics
Medical subspecialty
Others
No, 0
Mild, 1−2
Moderate, 3−4
Severe, >=5
Usual place of residence
Other hospital provider
Other places
0d
1d
2d
3d
4d
5d
6d
7−9d
10−13d
14−20d
21−27d
28+d
Alive
Died
F1
AUC
AUPRC
EO

**Figure S6. Additional model performance indicators by subgroups in the test dataset (01 February 2019 to 31 January 2020).** F1 score (a), area under the receiver operating curve (AUC) (b), and area under the precision-recall curve (AUPRC) (c) were compared. IMD=index of multiple deprivation score (higher scores indicate greater deprivation). Weekday refers to the day of the week of the index date. Source refers to the source of admission. Overall performance is shown by the dashed line in each plot. 95% confidence intervals were calculated using bootstrap. Balanced accuracy, positive predictive value (PPV), and negative predictive value (NPV) are shown in **Figure 3.** (d) Equalised odds (EO) differences for assessing model fairness, determined by either the per subgroup true positive rate or true negative rate differed from the overall rate by greater than an illustrative threshold of 0.1. 'O' represents a value ≤0.1 while 'X' represents a value >0.1.

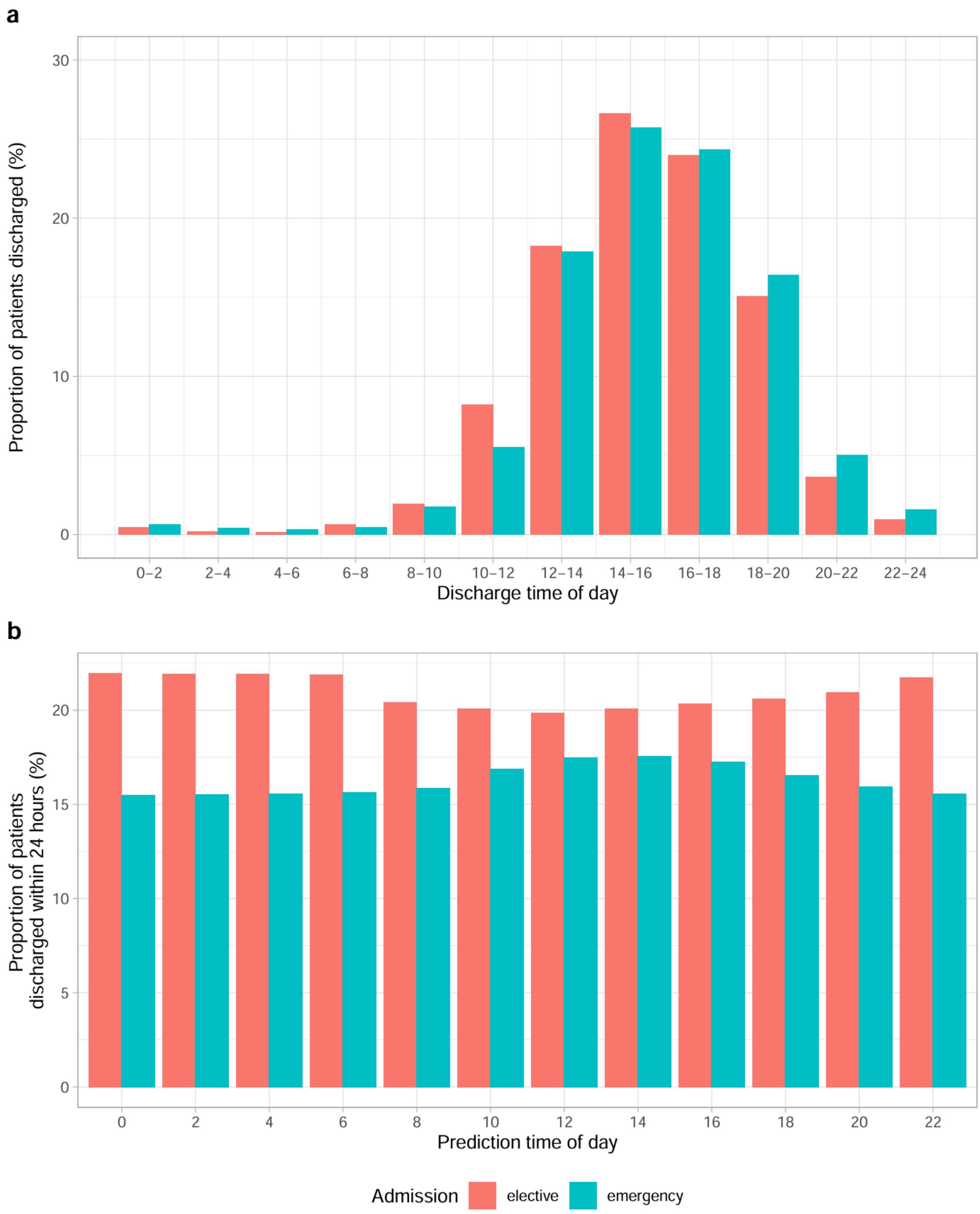

**Figure S7. Proportion of patients actually discharged by the observed hour of day of discharge (a) and proportion of patients discharged within the following 24 hours by model prediction time (b).**

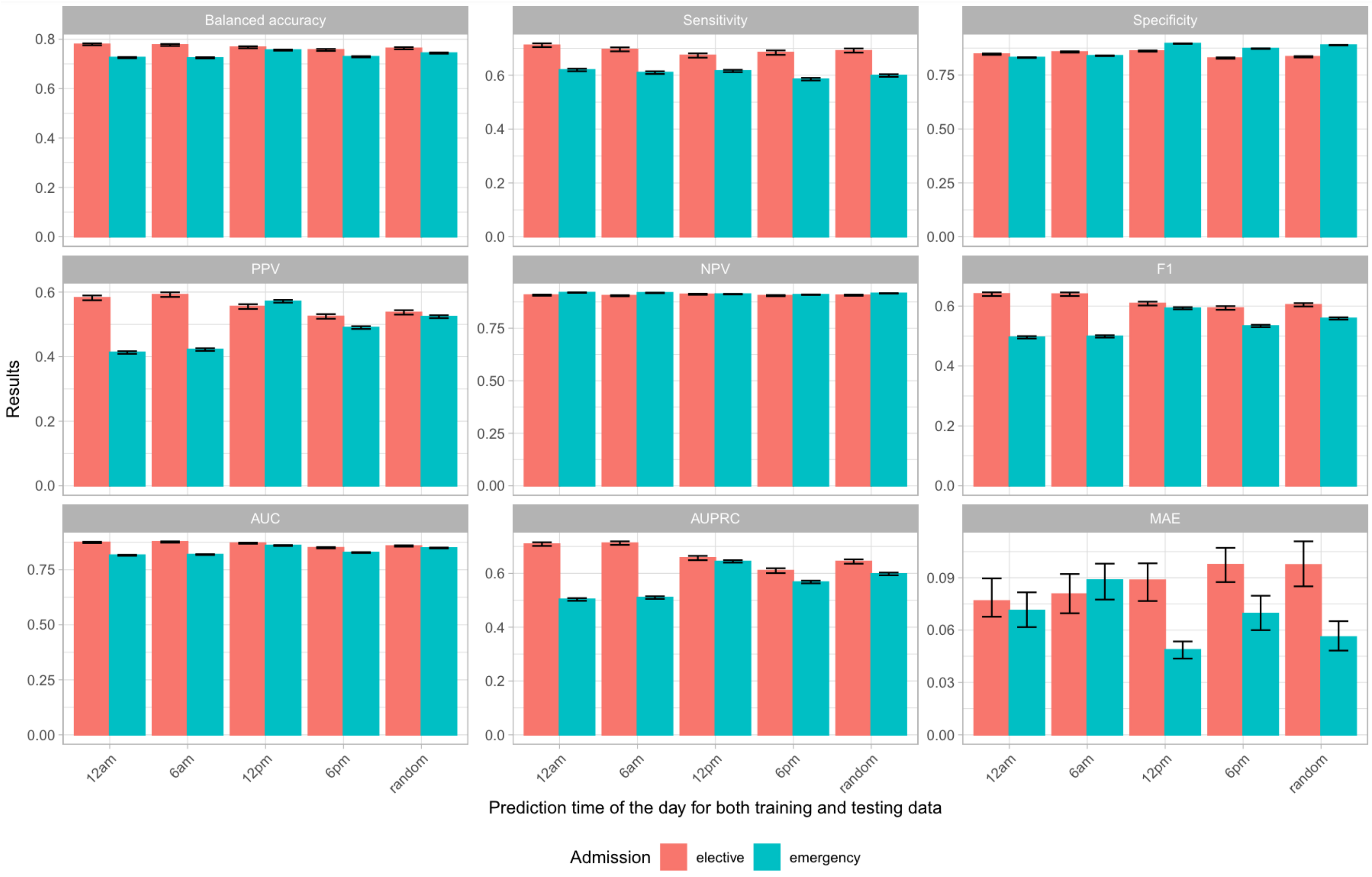

**Figure S8. Model performance using different prediction times of the day for elective and emergency admissions in the test dataset (01 February 2019 to 31 January 2020).** Predictions are shown for models trained at the same time of day. PPV: positive predictive value; NPV: negative predictive value; AUC: area under the receiver operating curve; AUPRC: area under the precision-recall curve; MAE: normalised mean absolute error (mean difference in predicted and actual discharges per day divided by the mean number of discharges per day).

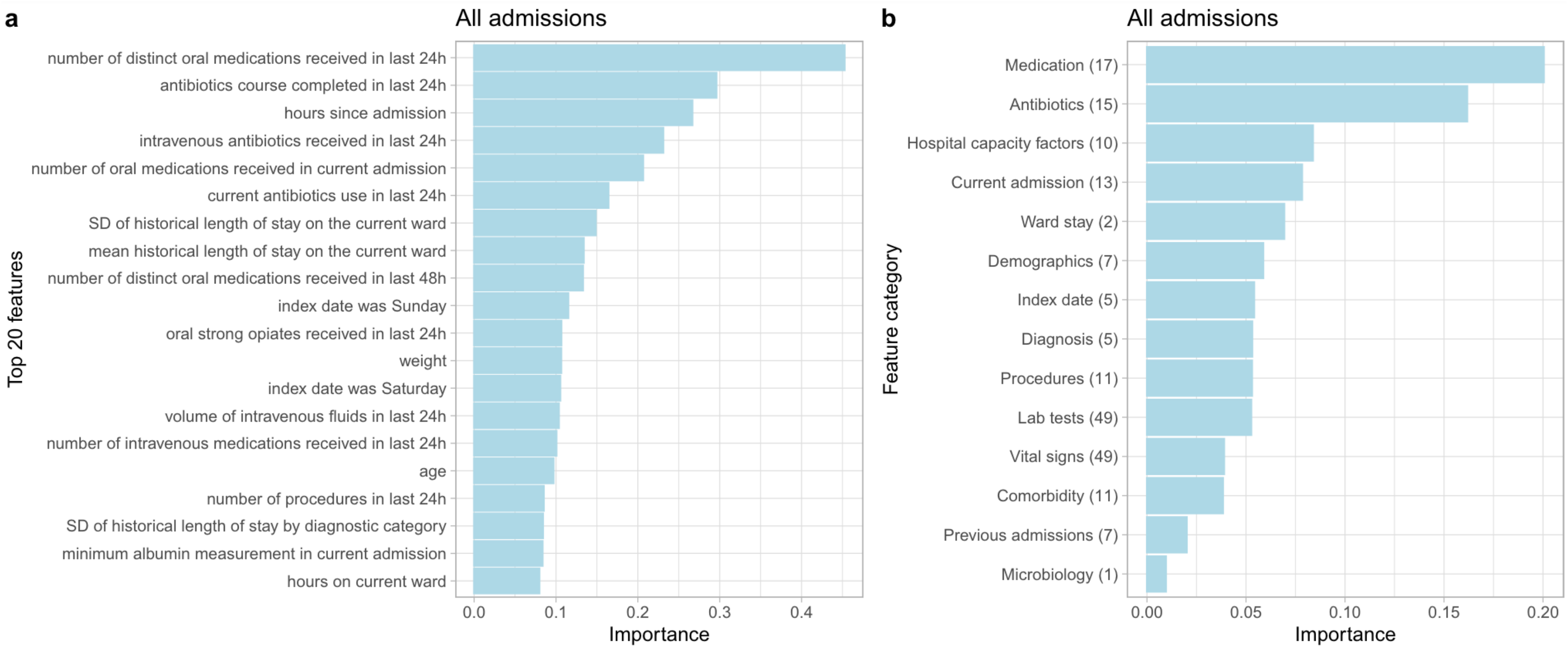

**Figure S9. Feature importance from the extreme gradient boosting model using SHAP values for all admissions, elective and emergency combined.** The top 20 most predictive features are shown in the order of predictiveness in panel **a**. Feature importance grouped by feature category is shown in the order of predictiveness in panel **b.** The mean importance of the top 5 most important features within each category is plotted. Numbers shown in parenthesis are number of features within the top 200 most predictive features in each category. No discharge planning features were selected. The complete list of features is summarised in **Table S1.** SHAP: SHapley Additive exPlanations. SD: standard deviation; LOS: length of stay; Current admission: admission time/source/specialty; SHMI: length of stay characteristics of SHMI diagnosis categories; Previous admissions: previous length of stay and readmission; Discharge: discharge planning.

| Category | Features | Number of features |
|---|---|---|
| Index date | ▪ Number of days since 1 January 2017<br>▪ Index date weekday<br>▪ Index date month<br>▪ Number of hours since admission | 4 |
| Demographics | ▪ Age<br>▪ Sex<br>▪ BMI, height, weight<br>▪ Ethnic group (white, mixed, Asian or Asian British, Black or Black British, other ethnic group, not stated/not known ethnic group)<br>▪ IMD score<br>▪ Prior mean, max, min, median, sd length of stay by postcode district | 13 |
| Comorbidities | ▪ Charlson comorbidity index, raw and age-adjusted[8]<br>▪ Elixhauser comorbidity score, raw and age-adjusted<br>▪ Individual comorbidities in Charlson score (acute myocardial infarction, cerebral vascular disease, congestive heart failure, connective tissue disorder, dementia, diabetes, liver disease, peptic ulcer, peripheral vascular disease, pulmonary disease, cancer, diabetes complications, paraplegia, renal disease, metastatic cancer, severe liver disease, HIV)<br>▪ Individual comorbidities in Elixhauser score (congestive heart failure, cardiac arrhythmias, valvular disease, pulmonary circulation disorders, peripheral vascular disorders, hypertension uncomplicated, paralysis, other neurological disorders chronic pulmonary disease, diabetes uncomplicated, diabetes complicated, hypothyroidism, renal failure, liver disease, peptic ulcer disease, HIV, lymphoma, metastatic cancer, solid tumour without metastasis, rheumatoid arthritis, coagulopathy, obesity, weight loss, fluid and electrolyte disorders, blood loss anaemia, deficiency anaemia, alcohol abuse, drug abuse, psychoses, depression, hypertension complicated)<br><br>All comorbidities are based on diagnostic codes in previous year before the current admission. | 52 |
| Current admission | ▪ Admission time/source features:<br>− daytime (0 to 24 hours)<br>− admission weekday (Monday to Sunday)<br>− admission month (January to December)<br>− admission source (usual place of residence, non-NHS institutional care, other NHS Provider)<br>▪ Admission specialty:<br>Acute, emergency and geriatric medicine; Acute and general surgery; Trauma and orthopaedics; Critical care; Medical subspecialty; Surgical subspecialty; Others.<br>− Specialty at index date<br>− Number of each specialty in current admission<br>− Number of unique specialties admitted within 365 days before the index date<br>− Number of new specialties under within last 24/48 hours | 7 |

| | | |
|---|---|---|
| Ward stay | ▪ Number of new wards within 24/48 hours<br>▪ The current ward is ICU<br>▪ Hours elapsed since the current ward starts | 4 |
| Diagnosis (Length of stay statistics for previous admissions for all patients, by SHMI category) | ▪ LOS characteristics of SHMI diagnosis categories: Historic mean/median/maximum/minimum/SD of the LOS of patients within the same SHMI diagnostic category | 8 |
| Discharge Planning | ▪ Physiotherapy referral within 24/48 hours before index date/ within 365 days before current admission date | 3 |
| Procedures | ▪ Had procedure within 24/48 hours before index date/within current admission<br>▪ Number of procedures 24/48 hours before index date/within current admission<br>▪ Time elapsed since most recent procedure before the index date, days<br>▪ Had procedure within 365 days before current admission date<br>▪ Number of procedures within 365 days before the current admission date | 21 |
| Antibiotics prescriptions | ▪ Current antibiotic use within 24/48 hours before the index date<br>▪ New antibiotics within 24/48 hours before the index date<br>▪ Antibiotics completed within 24/48 hours before index date<br>▪ Any antibiotics used within the current admission<br>▪ Duration of antibiotics used within current admission<br>▪ Count of unique antibiotics in the current admission<br>▪ Currently on a specific antibiotics agent (~60 antibiotics) | 73 |
| Medication | ▪ Use of intravenous fluids/intravenous medication/oral medication/nebulised medication/inhalation medication within 24/48 hours before index date/within current admission<br>▪ Volume of intravenous fluids within 24/48 hours before index date/within current admission<br>▪ Count of intravenous medication/oral medication/nebulised medication/inhalation medication within 24/48 hours before index date/within current admission<br>▪ Use of intravenous/oral strong opiates within 24/48 hours before index date/within current admission | 36 |
| Microbiology tests | ▪ Extended Spectrum Beta-Lactamase (ESBL)/ Carbapenemase-producing. Enterobacteriaceae (CPE) isolated in the current admission<br>▪ Vancomycin-resistant enterococci (VRE) isolated in the current admission<br>▪ Methicillin-resistant Staphylococcus aureus (MRSA) isolated in the current admission<br>▪ ESBL/CPE isolated in the last 365 days<br>▪ VRE isolated in last 365 days<br>▪ MRSA isolated in last 365 days<br>▪ Positive blood culture results within 24/48 hours before index date/within current admission | 6 |

| | | |
|---|---|---|
| Radiology investigation | ▪ Had radiology-based procedure within 24/48 hours before index date/within current admission<br>▪ Number of radiology-based procedures within 24/48 hours before index date/within current admission<br>▪ Had radiology-based procedure within 365 days before current admission date<br>▪ Number of radiology-based procedures within 365 days before the current admission date<br>▪ Time elapsed since most recent radiology procedure within 365 days before the index date | 9 |
| Readmissions and previous hospital stay | ▪ Readmissions<br>– Current admission is early readmission: ≤30 days from a previous hospitalization event<br>– Current admission is late readmission: >30 to ≤180 days from a previous hospitalization event<br>– Number of early 30-day readmissions within 365 days before the index date<br>– Time elapsed from most recent early 30-day readmission within 365 days before the index date, days<br>▪ Previous length of stay<br> – Number of admissions within 30/90/365 days before the index date<br> – Cumulative LOS within 30/90/365 days before the index date<br> – Mean/Maximum/Minimum/SD LOS per admission within 30/90/365 days before the current admission date | 22 |
| Hospital capacity factors | ▪ The median/mean/maximum/minimum/SD LOS of the current ward<br>▪ The current number of inpatients in the hospital<br>▪ Current inpatients with LOS to date of >= 7 days, 14 days, 28 days<br>▪ Proportion of current inpatients with LOS to date of >= 7 days, 14 days, 28 days<br>▪ Number of admissions within the last 24hr, 48hr, 7d, 28d<br>▪ Number of discharges within the last 24h, 48h, 7d, 28d<br>▪ The mean LOS for all discharges in the 7d, 14d, 28d before the index date | 23 |
| Vital signs | ▪ Mean / Max / Min / SD of each vital sign measurements (see below) within 24/48 hours before index date/within current admission<br>▪ Number of vital signs measurements within 24/48 hours before index date/within current admission<br><br>**List of vital signs:** heart rate, respiratory rate, systolic blood pressure, diastolic blood pressure, temperature, oxygen saturation, O2 L/min, O2 delivery device, AVPU score, NEWS2 score, NEWS2 score alternative (missing oxygen device = Room air) | 135 |
| Laboratory tests | ▪ Mean / Max / Min / SD of each laboratory test measurements within 24/48 hours before index date/within current admission<br>▪ Number of laboratory test measurements within 24/48 hours before index date/within current admission/within 365 days before the index date | 736 |

| | |
|---|---|
| | **List of laboratory tests:**<br>− **Complete blood counts:** Haemoglobin, Haematocrit, Mean Cell Volume, White Cell Count, Platelets, Neutrophils, Lymphocytes, Eosinophils, Monocytes, Basophils<br>− **Renal functions:** Creatinine, Urea, Potassium, Sodium, EGFR, Bicarbonate<br>− **Inflammatory:** C-reactive protein, Erythrocyte Sedimentation Rate, Creatinine kinase<br>− **Liver functions:** Alkaline phosphatase, Aspartate aminotransferase, Alanine transaminase, Albumin, Bilirubin, Amylase, Gamma-glutamyl Transferase<br>− **Bone/electrolytes profiles:** Adjusted calcium, Magnesium, Phosphate<br>− **Clotting:** Activated partial thromboplastin time, Prothrombin time, D_dimer,<br>− **Endocrine:** Thyroid-stimulating hormone, HbA1c, Glucose, Prostate-specific antigen<br>− **Haematinics:** Ferritin, Iron, Transferrin, B12, Folate<br>− **Others:** Lactate dehydrogenase, Troponin, Total Ig<br>− **Blood gases:** Base excess, Partial pressure of oxygen, Partial pressure of carbon dioxide, Lactate, Arterial blood pH<br>− **Lipids:** Triglycerides, High-density lipoprotein cholesterol, Total cholesterol, Low-density lipoprotein cholesterol |

**Table S1. Features included in the extreme gradient boosting model to predict hospital discharge within 24 hours.**

| Admission type | Data | Accuracy | Balanced accuracy | Sensitivity/Recall | Specificity | PPV/Precision | NPV | F1-score | AUC | AUPRC | MAE (%) |
|---|---|---|---|---|---|---|---|---|---|---|---|
| **Elective** | train | 0.930 | 0.938 | 0.951 | 0.926 | 0.757 | 0.987 | 0.843 | 0.981 | 0.918 | 4.3 |
| **Elective** | validation | 0.835 | 0.790 | 0.718 | 0.863 | 0.558 | 0.927 | 0.628 | 0.889 | 0.676 | 16.6 |
| **Elective** | test | 0.823 | 0.767 | 0.673 | 0.861 | 0.555 | 0.911 | 0.609 | 0.871 | 0.658 | 8.9 |
| **Emergency** | train | 0.901 | 0.842 | 0.753 | 0.932 | 0.693 | 0.948 | 0.721 | 0.950 | 0.792 | 3.6 |
| **Emergency** | validation | 0.854 | 0.757 | 0.609 | 0.904 | 0.566 | 0.918 | 0.587 | 0.866 | 0.638 | 8.0 |
| **Emergency** | test | 0.844 | 0.756 | 0.616 | 0.896 | 0.571 | 0.912 | 0.593 | 0.860 | 0.644 | 4.9 |
| **All** | train | 0.869 | 0.797 | 0.686 | 0.907 | 0.612 | 0.931 | 0.647 | 0.911 | 0.704 | 3.5 |
| **All** | validation | 0.846 | 0.765 | 0.639 | 0.891 | 0.561 | 0.919 | 0.597 | 0.872 | 0.643 | 7.2 |
| **All** | test | 0.837 | 0.752 | 0.615 | 0.888 | 0.561 | 0.909 | 0.587 | 0.858 | 0.634 | 4.6 |

**Table S2. Model performance of the extreme gradient boosting (XGB) model predicting 24-hour discharge in the training, validation, and test dataset.** PPV: positive predictive value; NPV: negative predictive value; AUC: area under the receiver operating curve; AUPRC: area under the precision-recall curve; MAE: normalised mean absolute error (mean difference in predicted and actual discharges per day divided by the mean number of discharges per day).

| Hyperparameter | Elective | Emergency | Overall | Optimisation options |
|---|---|---|---|---|
| **colsample_bytree** | 0.5 | 0.8 | 0.4 | 0.2, 0.3, 0.4, 0.5, 0.6, 0.7, 0.8, 0.9, 1 |
| **gamma** | 0 | 0.3 | 0.4 | 0.1, 0.2, 0.3, 0.4, 0.5 |
| **learning_rate** | 0.05 | 0.05 | 0.2 | 0.001, 0.005, 0.01, 0.05, 0.1, 0.2, 0.5, 1 |
| **max_depth** | 7 | 7 | 6 | 3, 4, 5, 6, 7, 8, 9, 10, 11, 12 |
| **min_child_weight** | 2 | 3 | 2 | 1, 2, 3, 4, 5, 6 |
| **n_estimators** | 775 | 750 | 400 | 50, 75, 100, 125, …, 1200 |
| **reg_alpha** | 0.01 | 0.01 | 0.0001 | 0.0001, 0.001, 0.01, 0.1, 1, 10, 100 |
| **reg_lambda** | 0.01 | 10 | 100 | 0.0001, 0.001, 0.01, 0.1, 1, 10, 100 |
| **subsample** | 0.9 | 0.9 | 0.9 | 0.3, 0.4, 0.5, 0.6, 0.7, 0.8, 0.9, 1 |

**Table S3. Hyperparameters chosen by the extreme gradient boosting models.**
